# The Tumor Profiler Study: Integrated, multi-omic, functional tumor profiling for clinical decision support

**DOI:** 10.1101/2020.02.13.20017921

**Authors:** Anja Irmisch, Ximena Bonilla, Stéphane Chevrier, Kjong-Van Lehmann, Franziska Singer, Nora C Toussaint, Cinzia Esposito, Julien Mena, Emanuela S Milani, Ruben Casanova, Daniel J Stekhoven, Rebekka Wegmann, Francis Jacob, Bettina Sobottka, Sandra Goetze, Jack Kuipers, Jacobo Sarabia del Castillo, Michael Prummer, Mustafa Tuncel, Ulrike Menzel, Andrea Jacobs, Stefanie Engler, Sujana Sivapatham, Anja Frei, Gabriele Gut, Joanna Ficek, Reinhard Dummer, Tumor Profiler Consortium, Rudolf Aebersold, Marina Bacac, Niko Beerenwinkel, Christian Beisel, Bernd Bodenmiller, Viktor H Koelzer, Holger Moch, Lucas Pelkmans, Berend Snijder, Markus Tolnay, Bernd Wollscheid, Gunnar Rätsch, Mitchell Levesque

## Abstract

Recent technological advances allow profiling of tumor samples to an unparalleled level with respect to molecular and spatial composition as well as treatment response. We describe a prospective, observational clinical study performed within the Tumor Profiler (TuPro) Consortium that aims to show the extent to which such comprehensive information leads to advanced mechanistic insights of a patient’s tumor, enables prognostic and predictive biomarker discovery, and has the potential to support clinical decision making. For this study of melanoma, ovarian carcinoma, and acute myeloid leukemia tumors, in addition to the emerging standard diagnostic approaches of targeted NGS panel sequencing and digital pathology, we perform extensive characterization using the following exploratory technologies: single-cell genomics and transcriptomics, proteotyping, CyTOF, imaging CyTOF, pharmacoscopy, and 4i drug response profiling (4i DRP). In this work, we outline the aims of the TuPro study and present preliminary results on the feasibility of using these technologies in clinical practice showcasing the power of an integrative multi-modal and functional approach for understanding a tumor’s underlying biology and for clinical decision support.

## Introduction

In recent years, the development of numerous drugs targeting oncogenic pathways and immune checkpoints, as well as the advent of next-generation sequencing (NGS) and its continuous improvement, higher availability, and decreasing cost, have allowed major cancer centers worldwide to offer NGS-based personalized treatment options for clinical practice (Beltran et al. 2015; Massard et al. 2017; Meric-Bernstam et al. 2015; Le Tourneau et al. 2015). In those studies, treatments are suggested according to the tumor’s genetic profile and are agnostic to the tissue of origin. These NGS-focused approaches have had some successes so far, with about one-third of patients treated with NGS-matched treatments showing a significant tumor response (Massard et al. 2017; Rodon et al. 2019; Sicklick et al. 2019; F. Singer et al. 2018). However, since NGS-based approaches typically only analyze genomic alterations from bulk tissue, many clinically-relevant questions about the tumor, such as its micro-environment, heterogeneity, and many others, remain unanswered (Friedman et al. 2015). In this study, we explore to which extent a broader and more in-depth functional profiling of tumors can lead to an enhanced understanding of the biology of a tumor and whether this new knowledge is actionable for the treatment of cancer patients.

Technological progress has made the precise quantification of the molecular make-up of cells, their biochemical state, their interactions in tumor tissue, and their *ex vivo* response to drugs increasingly feasible. To take advantage of these recent advances, the Tumor Profiler (TuPro) consortium was formed as a collaborative effort to combine exploratory technologies that have been pioneered in the participating academic institutions. The goal is to decipher the in-depth molecular and functional landscape of patient tumors across different indications, namely melanoma, ovarian carcinoma, and acute myeloid leukemia (AML), to set up a platform for multi-omics clinical decision support in an observational clinical study. As part of the TuPro workflow, changes at the DNA, RNA, and protein level are assessed at single-cell resolution in order to characterize both the tumor and the immune compartment. Samples are also analyzed at the whole-tissue level with targeted NGS, proteotyping, and bulk RNA sequencing. Moreover, *ex vivo* drug response testing is performed to assess the effect of selected drugs on cancer cells. Digital pathology based on large-scale immunohistochemistry analysis, as well as in-depth highly multiplexed imaging analysis, is included to provide spatially resolved data. These technologies are unique in combination, provide unprecedented insights into the molecular, cellular, and functional state of human tumor tissues, and form the technical basis of the study that is presented here.

Initial TuPro results show that a clinically relevant turnaround time of two weeks is feasible for all the technologies included in the trial, that analysis results are consistent between technologies, and that integrated results provide treatment decision support beyond an approved diagnostic NGS panel sequencing.

## Study Design

### Concept

The TuPro study is designed as an observational clinical study in which patients provide full informed consent (BASEC-Nr.2018-02050, approved by the Kantonal Ethics Commisions of Zürich and Basel) to have their tumors analyzed by a compendium of advanced technologies. The goal is to ultimately assess whether a combination of multi-omics and functional profiling techniques (see “Technologies”) on the same tumor tissue can supplement emerging standards, such as digital pathology and targeted NGS, to support molecularly informed clinical decision making in tumor boards.

The technologies included in the trial were selected based on their ability to explore a tumor in multiple ways at different molecular and cellular levels, from bulk to single-cell assays, targeted to non-targeted approaches, RNA to protein expression, cells in suspension to spatially-resolved tissues, and phenotypic to functional readouts. The TuPro study aims to determine how each technology contributes to the identification of clinically relevant features in a sample and how the integration of all data types could potentially improve treatment decisions for each patient enrolled in the trial. Ultimately, the synergistic effect of this multi-modal approach is expected to enable the establishment of algorithms for integrative analyses and to provide relevant information that can be used to inform patient management directly.

Approximately 1cm^3^ of solid tumor material from patients fulfilling the inclusion criteria (see “Indication” below) is distributed to all the technology platforms, which analyze the sample and return their data within two weeks. These data are then integrated into a comprehensive molecular research report (MRR). This report is used in a pre-Tumor Board (pre-TB) discussion where hypothetical treatment decisions based on TuPro data are compared to treatment decisions that would have been taken based on a) standard clinical guidelines (ESMO clinical guidelines (Dummer et al. 2015; Colombo et al. 2019; Fey, Buske, and ESMO Guidelines Working Group 2013) and b) emerging clinical practice that includes targeted NGS. Up to this point, TuPro R&D has focused on the “Fast Diagnostics Loop” in which patients are recruited, and the pre-TB receives treatment recommendations within four weeks after biopsy (**Figure 1A**). A summary of the TuPro MRR, including the hypothetical treatment recommendations of the pre-TB, is communicated to the Tumor Board (TB). This expert panel makes the final decision on the best treatment strategy, given the available information on the individual patient. Clinical follow-up data, such as performance status (ECOG, see below), treatment, side effects, and response data from all TuPro patients are recorded and analysed in conjunction with the molecular data provided by the TuPro technologies within an “Exploratory Science Loop”, where hypothesis-generating analyses are carried out during and at the end of the clinical study (**Figure 1A**).

**Figure 1:**
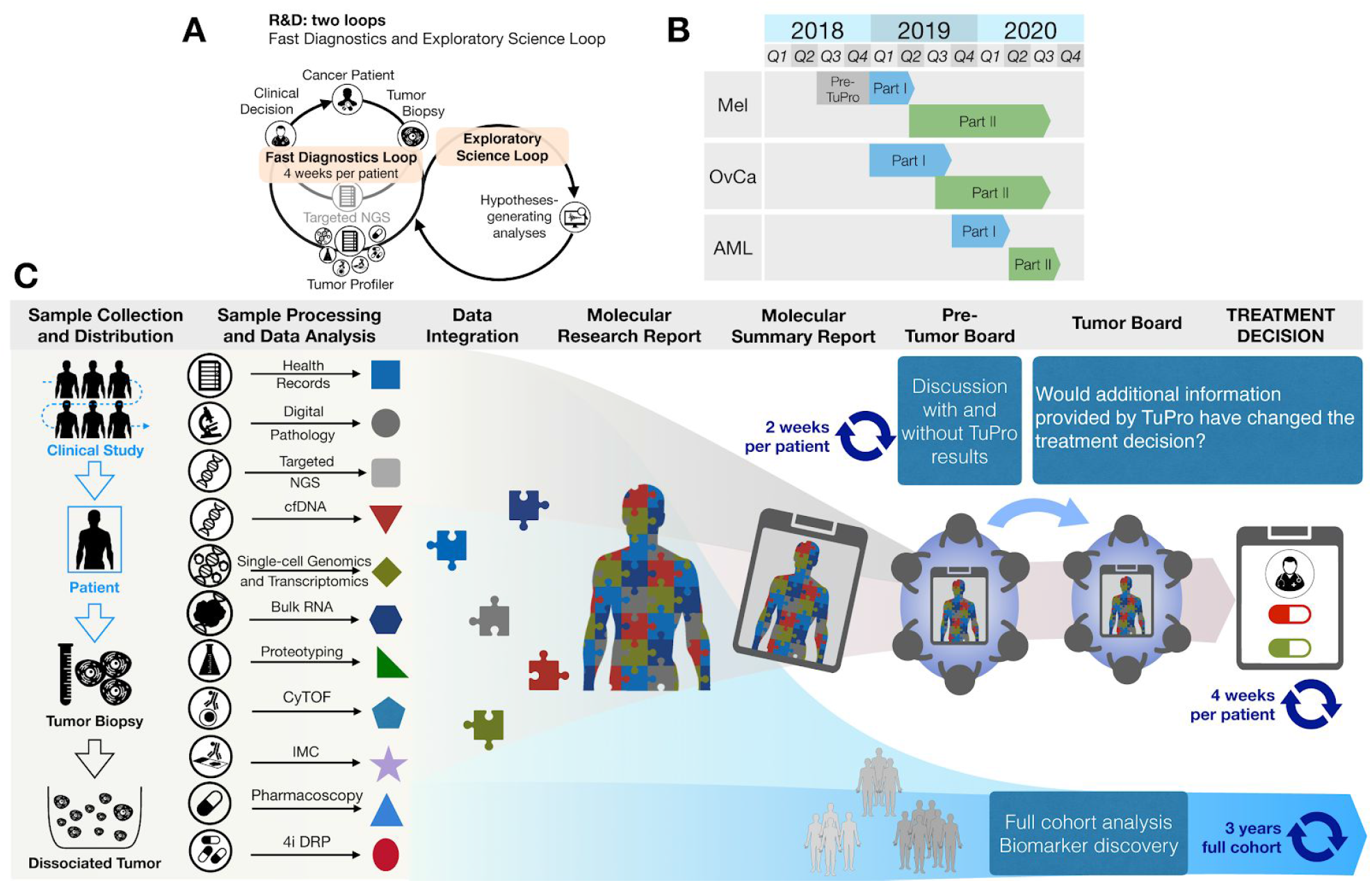
Overview of the Tumor Profiler Study. **(A)** The R&D workstream consists of two loops, the Fast Diagnostics Loop, which provides integrated information from approved diagnostics and additional TuPro exploratory technologies in a four week turnaround time, and the Exploratory Science Loop for which cohort analysis is performed during and at the end of the clinical study. **(B)** Tumor Profiler includes three indications: malignant melanoma (Mel), ovarian carcinoma (OvCa), and acute myeloid leukemia (AML). The Pre-TuPro consisted of a pre-proof-of-concept phase; Part I comprised the proof-of-concept phase; Part II the study phase. **(C)** Detailed overview of patient enrollment, sample collection, analysis by different technology platforms, data integration, creation and discussion of molecular research and summary reports, clinical discussion in pre-TBs which result in hypothetical treatment decisions for each patient based on routine diagnostics alone and after consideration of TuPro data, and Tumor Boards.

### Consortium Structure

TuPro is a cooperative, multi-institutional project involving F. Hoffmann-La Roche AG (Roche), ETH Zurich, University of Zurich, University Hospital Zurich, and University Hospital Basel (consortium author list in the Supplementary Material). These institutions provide the expertise for the establishment of the three workstreams required for the TuPro initiative, including the clinical, R&D, and data/IT workstreams. The clinical workstream is responsible for patient screening and selection, and for providing histopathological analysis. The R&D workstream investigates the molecular make-up of tumors based on six cutting-edge technologies and performs integrative analyses (see Section “Technologies” below). The data/IT workstream provides the data and computational infrastructure and manages data access for all partners. It interacts as well with the clinical workstream to produce a comprehensive and integrative report of the findings of the technology nodes that is presented to the pre-TB.

### Indications

The TuPro study includes patients with three different indications: melanoma, ovarian carcinoma, and acute myeloid leukemia (AML). For melanoma, any patients with stage III/IV cutaneous melanoma can be included; patients with rare melanomas, such as mucosal and ocular melanomas, can be recruited independently of the stage, provided that they require systemic (including adjuvant) therapy. For ovarian carcinoma, patients with high-grade primary or recurrent adenocarcinoma of ovarian, tubal, or peritoneal origin at FIGO stage III/IV can be recruited. For AML, patients with relapsed/refractory AML can be included. For both melanoma and ovarian carcinoma, a minimum ECOG of 2 is required for the patient to enter the study. In total, up to 240 patients will be included in the study across the three indications.

### Study Timeline

The TuPro project is based on the sequential inclusion of different tumor indications, and clinical studies for all indications will run until the end of 2020 (last patient, last visit). Recruitment of melanoma and ovarian carcinoma started in December 2018 and January 2019, respectively. Recruitment for AML started in September 2019 **(Figure 1B)**.

The selection of each of the TuPro indications has been preceded by an evaluation phase during which the clinical needs, the scientific relevance, and the feasibility of the inclusion of the respective indication have been investigated. Once the relevance of a specific indication has been established, samples corresponding to the approved indications are included in the study. For each indication, the study consists of two parts. The first part aims to refine and standardize the experimental and analytical processes within the different workstreams. During this phase, the data generated by the different technologies are not taken into account when making treatment decisions, and the results are not discussed with the patients. During the second part of the project, molecular summary reports summarizing the findings of the TuPro technologies and the pre-TB treatment recommendations are shared with the Tumor Board. The Tumor Board then makes a recommendation based on all the available data. The final decision on which treatment to implement is recorded.

### Technology Platforms

The main focus of TuPro is to assess the feasibility and impact of new molecular and cellular profiling technologies and the integration of the respective data to gain a more complex understanding of a tumor and, if possible, on the pharmacological treatment of the patients enrolled in the study. A second important goal is to generate a dataset suitable to discover molecular and digital biomarkers relevant for treatment recommendations **(Figure 1C)**. We outline below the profiling technologies included in this study (**Table 1**).

**Table 1:**
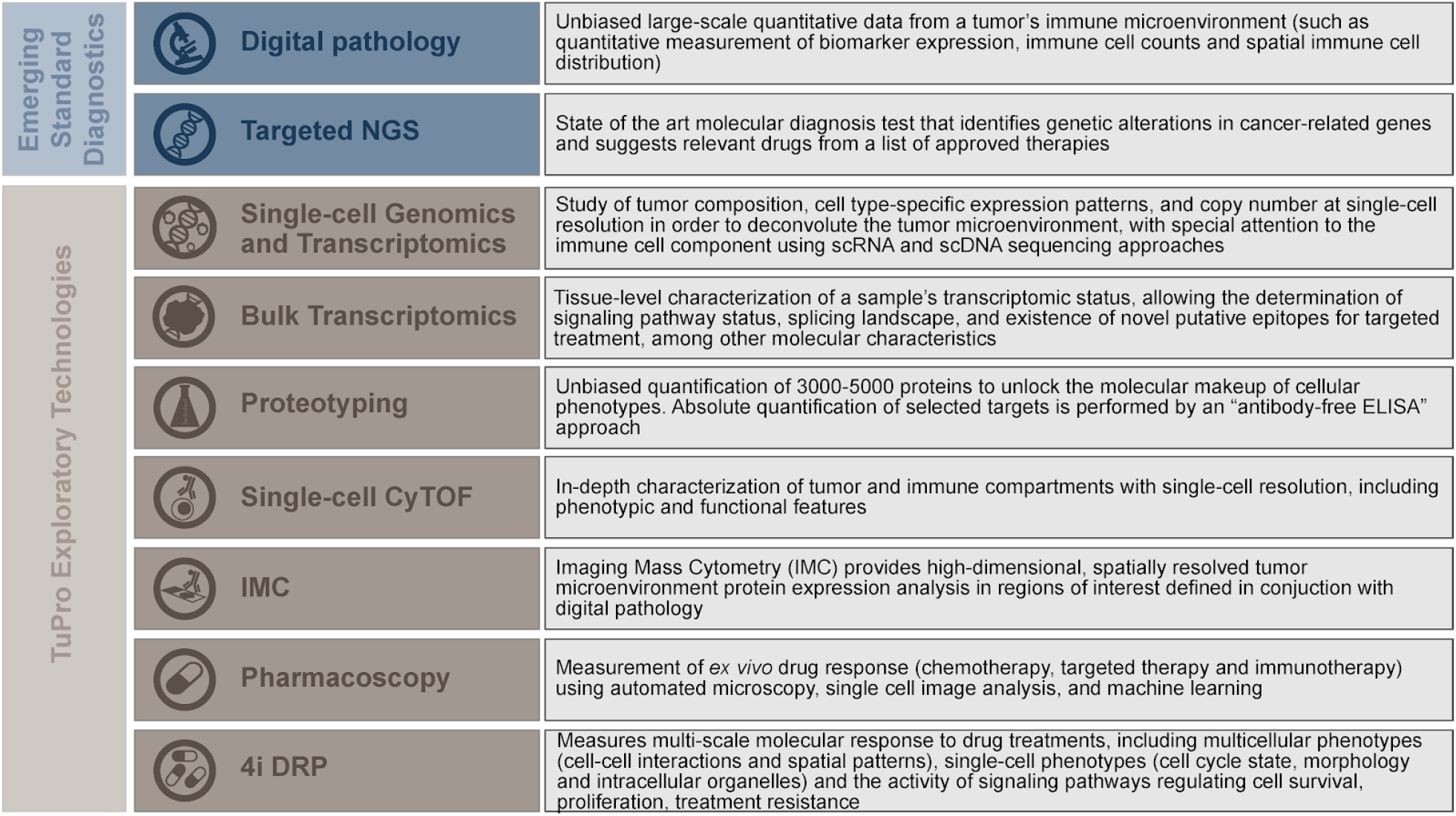
Molecular profiling technologies used in TuPro. The molecular approach utilized and the type of data the technologies included in TuPro contribute to the study are briefly summarized in this table. The left-most panel groups the technologies within two categories based on the level of clinical-diagnosis approval (Emerging Standard Diagnostics and TuPro Exploratory Technologies).

|  |  |  |
| --- | --- | --- |
| Emerging Standard Diagnostics | **Digital pathology** | Unbiased large-scale quantitative data from a tumor's immune microenvironment (such as quantitative measurement of biomarker expression, immune cell counts and spatial immune cell distribution) |
|  | **Targeted NGS** | State of the art molecular diagnosis test that identifies genetic alterations in cancer-related genes and suggests relevant drugs from a list of approved therapies |
| TuPro Exploratory Technologies | **Single-cell Genomics and Transcriptomics** | Study of tumor composition, cell type-specific expression patterns, and copy number at single-cell resolution in order to deconvolute the tumor microenvironment, with special attention to the immune cell component using scRNA and scDNA sequencing approaches |
|  | **Bulk Transcriptomics** | Tissue-level characterization of a sample's transcriptomic status, allowing the determination of signaling pathway status, splicing landscape, and existence of novel putative epitopes for targeted treatment, among other molecular characteristics |
|  | **Proteotyping** | Unbiased quantification of 3000-5000 proteins to unlock the molecular makeup of cellular phenotypes. Absolute quantification of selected targets is performed by an "antibody-free ELISA" approach |
|  | **Single-cell CyTOF** | In-depth characterization of tumor and immune compartments with single-cell resolution, including phenotypic and functional features |
|  | **IMC** | Imaging Mass Cytometry (IMC) provides high-dimensional, spatially resolved tumor microenvironment protein expression analysis in regions of interest defined in conjunction with digital pathology |
|  | **Pharmacoscopy** | Measurement of *ex vivo* drug response (chemotherapy, targeted therapy and immunotherapy) using automated microscopy, single cell image analysis, and machine learning |
|  | **4i DRP** | Measures multi-scale molecular response to drug treatments, including multicellular phenotypes (cell-cell interactions and spatial patterns), single-cell phenotypes (cell cycle state, morphology and intracellular organelles) and the activity of signaling pathways regulating cell survival, proliferation, treatment resistance |

#### Sample preparation and pathology diagnosis

A pathologist reviews every TuPro tissue sample for suitability and selection of vital tumor tissue for the technology platforms. Pathology diagnosis includes the confirmation of tumor type by conventional pathology as well as immunohistochemical staining (IHC) for tumor-specific biomarkers (e.g., S100, SOX10, and MelanA for melanoma and p53, ER, PR, PAX8, WT1, NAPSIN, and CDX2 for ovarian carcinomas) and assessment of tumor cell content. Only samples with a definite tissue diagnosis and limited necrotic tissue are included in the TuPro trial. For melanoma, we additionally require a minimum of 20% vital tumor cells. Fresh tissue samples are released for single-cell dissociation and subsequent distribution to the TuPro technologies while FFPE sections are sent to digital pathology, targeted NGS, proteotyping, and imaging mass cytometry (IMC).

### Emerging Standard Diagnostics

#### Digital Pathology

Immune phenotyping is performed by IHC for CD3+ pan T cells, CD4+ T-helper cells, CD8+ cytotoxic T cells, Foxp3+ regulatory T cells as well as CD20+ B-cells and CD68+ macrophage populations. Expression of the immune checkpoints PD1 and PD-L1 is quantified, and specific features of the immune microenvironment (e.g., the formation of tertiary lymphoid structures) are recorded. Based on the quantity and distribution of the immune markers, tumors are classified as “inflamed” (strong intra- and peritumoral immune cell infiltration), “immune excluded” (immune response around the tumor but no infiltration into tumor cell clusters) or “immune desert” (absence of immune response) to support treatment decisions in conjunction with the TuPro technology nodes. Digital image analysis is applied to provide unbiased, large-scale quantitative data for the core immune markers on all TuPro samples. This includes absolute quantification of each cell population as well as spatial information to account for tumor heterogeneity and to enable tumor infiltration analyses. TuPro digital pathology images are then used to select the optimal regions for highly multiplexed assessment by imaging mass cytometry.

#### Targeted Next-Generation Sequencing

Targeted Next-Generation Sequencing (NGS) was performed with the *FDA*-approved, broad comprehensive molecular diagnostic test FoundationOne^®^CDx assay (Foundation Medicine Inc., Cambridge, MA, USA). The assay sequences the complete exons of 324 cancer-related genes for the detection of short variants (single nucleotide variants, insertions, and deletions), copy number alterations (CNAs), as well as microsatellite status and tumor mutational burden. Selected introns and promoter regions of 36 genes are also sequenced for the detection of gene rearrangements and fusions (Frampton et al. 2013). A report containing clinically actionable variants, along with treatment recommendations suggested by Foundation Medicine, Inc. proprietary algorithms, is provided to the TuPro consortium for integration in their analyses.

To complement the targeted NGS analyses, we investigate circulating tumor DNA (ctDNA, **Supplementary note 1**) as a minimally invasive tool to monitor disease progression and recurrence (Bettegowda et al. 2014).

### TuPro Exploratory Technologies

#### Cell Types, Markers, and Drug Selection

Two types of clinically relevant markers are analyzed by TuPro technologies. First, markers that allow a detailed determination of the composition of the tumor and its microenvironment, and second, predictive markers for clinical decision support. These marker sets are selected for each indication separately. For more details on marker selection, please refer to **Supplementary Note 2**.

#### Single-cell Genomics and Transcriptomics

The Single-Cell Genomics and Transcriptomics platform (**Supplementary Figure 1**) uses droplet-based scDNA-seq and scRNA-seq protocols to identify copy number changes in tumor subclones and their evolutionary relationships, to quantify different cell populations, and to elucidate their gene expression patterns (Hedlund and Deng 2018; Tirosh et al. 2016; Shih et al. 2018; Svensson, Vento-Tormo, and Teichmann 2018; Zahn et al. 2017). This approach probes the cellular heterogeneity of the tumor and its local environment and the molecular phenotype in an untargeted and genome-wide fashion on the level of individual cells. Through modular and flexible computational data analysis pipelines, it provides a detailed picture of the tumor and its microenvironment (Tirosh et al. 2016), as well as its phylogenetic structure (Navin et al. 2011; Jahn, Kuipers, and Beerenwinkel 2016; Kuipers et al. 2017; J. Singer et al. 2018). The genomic and transcriptomic readouts reveal the distribution of tumor marker genes, drug targets, and immunoregulatory markers across cells (Ho et al. 2018). Gene expression profiles of subclones inform on aberrant gene sets and are analyzed further for pathway enrichment and for subclone-specific *in-silico* drug predictions. The presence of different immune cell types and their expression profiles enlightens the interplay between tumor and non-malignant cells and has direct implications for the effectiveness of immunotherapies (Papalexi and Satija 2018; Paulson et al. 2018). Assessing the genome and transcriptome of complex tumors and their microenvironment may help in the characterization of the driving forces of tumor evolution and guide therapies to become more successful in treating the whole tumor ecosystem.

#### Bulk Transcriptomics

To obtain information about alternative splicing, alternative promoters, gene-fusions, RNA-editing and other RNA aberrations (Kahles et al. 2018; PCAWG Transcriptome Core Group et al. 2018) and to complement the other transcriptome and proteome profiling technologies, we also obtain bulk-RNA-seq data, when sufficient tumor material is available. Also, the additional RNA-seq data from the samples in this study provides a common basis for cross-comparison and joint analyses with samples from TCGA, ICGC, GTEx and other studies (Cancer Genome Atlas Network 2015; Campbell et al. 2017; The GTEx Consortium 2015).

#### Proteotype Analysis

The clinical proteotyping platform enables the conversion of a clinical sample into a permanent digital proteotype map. This digital proteotype map contains information about the state of the proteome at the time of the measurement (“proteotype”), which includes the identity and quantity of peptides/proteins, as well as their post-translational modifications (Röst, Malmström, and Aebersold 2015; Aebersold and Mann 2016). Such digital proteotype maps derived from clinical specimens can be analyzed, re-analyzed, compared, and mined *in silico* to detect and quantify peptide/protein patterns across large clinical cohorts (Guo et al. 2015). Within the context of the TuPro study Data-Independent-Acquisition Mass Spectrometry (DIA-MS) (Gillet et al. 2012; Bruderer et al. 2015) and targeted mass spectrometry (PRM, parallel reaction monitoring) (Peterson et al. 2012; Schiess, Wollscheid, and Aebersold 2009) are used to generate such digital proteotype maps. Phenotyping cells/tissues without the need for antibodies using DIA-MS and PRM provides information about the abundance level of peptides/proteins within cells/tissues which are current clinical drug targets and reveals protein abundance patterns which can be correlated across cohort samples from clinical trials for drug response prediction (**Supplementary Figure 2**).

#### Mass Cytometry (CyTOF)

Mass cytometry allows for the simultaneous quantification of over 50 proteins, phosphorylation sites, and transcripts on the single-cell level based on the use of metal-labeled reporters (Bandura et al. 2009; Frei et al. 2016). With its high dynamic range, low detection limit, high-throughput capability and relatively low cost, this method has proven to be particularly well-suited for the analysis of highly heterogeneous tumor samples (Chevrier et al. 2017; Lavin et al. 2017; Wagner et al. 2019). In the context of the TuPro study, mass cytometry analysis is based on two 40-antibody panels per indication, designed to characterize the immune and the tumor compartment of each sample, respectively, with a specific emphasis on markers which can be targeted in immuno- and onco-therapies (**Supplementary Figure 3**).

#### Imaging Mass Cytometry (IMC)

IMC allows for the quantification of over 50 markers with subcellular resolution on a single tissue section (Giesen et al. 2014; Schapiro et al. 2017; Schulz et al. 2018). This enables the identification of tissue morphology and structural features such as vessels, stroma, tumor tissue phenotypes, and a variety of immune cell types on selected regions of the different tumor samples. IMC provides high dimensional single-cell phenotyping as its main readout, thus allowing quantification of the proportions of immune and tumor cell populations. Based on a 40-antibody panel per indication developed specifically for the TuPro study, information on spatial interactions between cell populations with a focus on tumor-immune cell interactions is generated which helps in the assessment of immune infiltration patterns (**Supplementary Figure 4**). This technology also enables the identification of the direct neighbors of a specific cell type, including for instance physical interactions between PD-1 positive T cells and PD-L1 expressing tumor or myeloid cells (Giesen et al. 2014; Schapiro et al. 2017; Damond et al. 2019).

#### Pharmacoscopy

The pharmacoscopy node uses high-throughput microscopy to measure *ex vivo* responses to targeted therapies, chemotherapies, and immunotherapies. The method provides a drug sensitivity ranking for a large number of drugs and combinations using a functional relative toxicity measurement of the cancer cells against healthy cells within the sample. The readout can be directly used to identify effective therapies for individual patients as well as to improve the understanding of the cellular and molecular systems that determine drug response variability (**Supplementary Figure 5**). Clinical benefits were previously demonstrated in a study with hematological malignancies (Snijder et al. 2017).

#### 4i Drug Response Profiling (4i DRP)

The 4i Drug Response Profiling (4i DRP) technology utilizes an innovative multiplexed immunofluorescence protocol, termed 4i (iterative indirect immunofluorescence imaging) (Gut, Herrmann, and Pelkmans 2018), to achieve an in-depth molecular characterization of phenotypic patient responses to a broad spectrum of drug categories (**Supplementary Figure 6**). Implemented with large-scale image processing, computer vision, and machine learning, the 4i technology is a modern approach in precision medicine to uncover activation of key proliferation and survival pathways as well as the emergence of drug resistance in tumor biopsies profiled with a dedicated drug library, at single-cell resolution. The multivariate analysis of multiplexed regulatory signaling proteins in individual cells and the associated multiscale cellular state (morphology, cell size, cell cycle, etc.) allows to decipher cell-to-cell phenotypic variability to different drug treatments and generate predictive models of drug response. This is supported by evidence that cell heterogeneity in steady-state or in perturbed state can be predicted by a multivariate set of cell features (Snijder et al. 2009; Battich, Stoeger, and Pelkmans 2015). On a larger cohort of patients, this approach may contribute to the identification of predictive and/or prognostic biomarkers and the development of new targeted therapies.

### Data Analysis, Reports, and Tumor boards

The clinical and molecular data are collected, stored, and analyzed in a research data management system (RDMS) developed for this project (**Supplementary Figure 7**).

Based on the collected data and technology-specific analyses, a multidisciplinary team consisting of technology experts, computational biologists, pathologists, and clinical experts jointly generates the Molecular Research Report (MRR) (**Figure 3**). The MRR is accessible via a web application intended to be used in the pre-TB, where it facilitates discussions between technology experts and clinicians. It is structured in two parts: the R&D platform pages and the front page.

**Figure 3:**
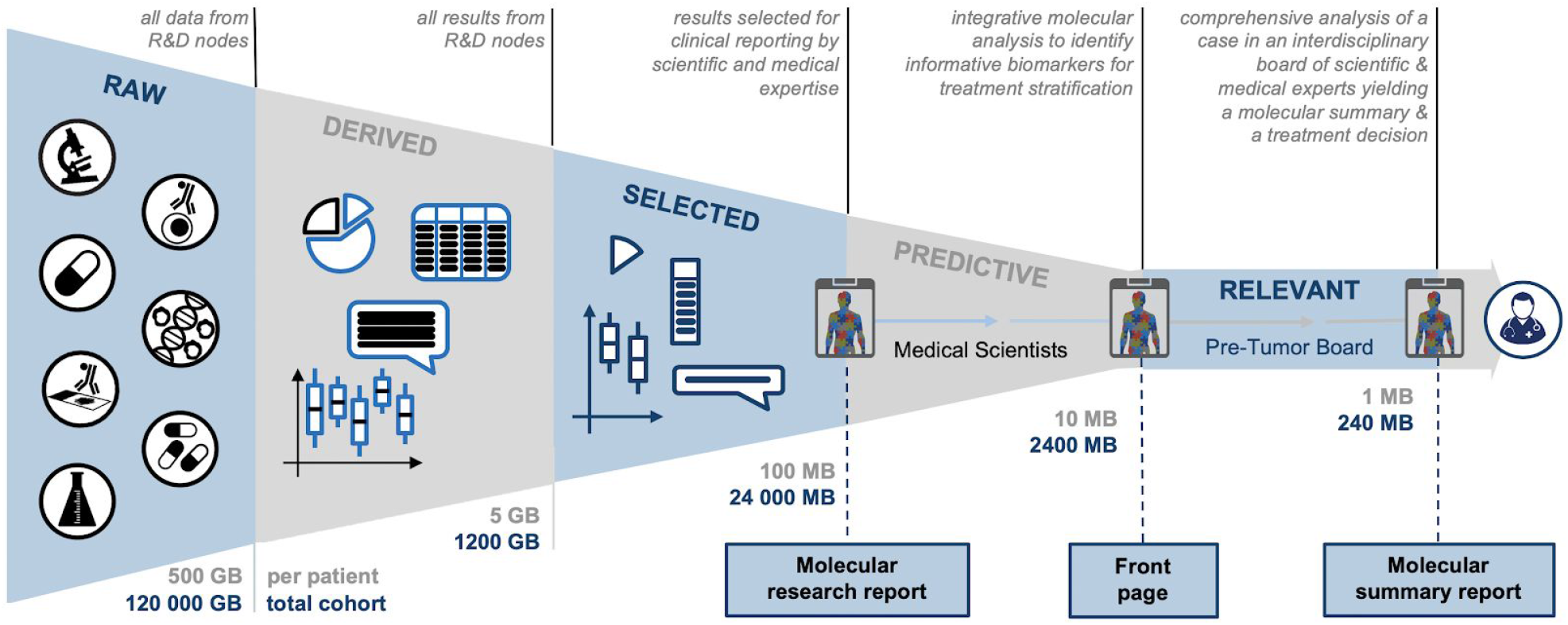
Schematic representation of the qualitative and quantitative transition from the raw data generated by all R&D nodes (left) to the molecular summary report (MSR, right). For each patient, the R&D nodes generate up to 500 GB of raw data. The derived results represent a comprehensive molecular profile of each patient. Based on medical and scientific domain expertise, a subset of derived results is selected for reporting and thus forms the basis for the molecular research report (MRR). Integrating the different modalities with patient data and existing knowledge, predictive biomarkers are summarized on the front page of the MRR. This serves as the basis for the selection of a potential treatment in the pre-TB, which ultimately is composing the MSR, which is then discussed by the Tumor Board deciding on the treatment.

The **R&D platform pages** present the analysis results from the individual R&D platforms separately, using dynamic figures and tables. Clinically relevant results are summarized at the top of the page. The R&D platform pages are considered to be comprehensive for each processed sample, providing a full, yet unimodal patient-level representation from the respective technology.

The **front page** of the MRR is an integrative summary of the clinically relevant R&D results with a focus on the prioritization of treatment options. The MRR front page is structured in three parts: 1) patient information, e.g., unique ID, gender, age-range, disease stage; 2) histological and molecular phenotype, and 3) overview of potential treatments. It provides a concise overview of potential treatments and corresponding supporting evidence from all platforms.

The goal of reporting within the TuPro study is not only to present the results in a meaningful way, but also to document the pre-TB’s process leading to a specific treatment recommendation. Therefore, after the pre-TB, a summary of the discussion and the treatment recommendation are added to the MRR front page. The pre-TB treatment recommendation is recorded first based on the standard treatment guidelines and based on emerging standard diagnostics and again after the presentation of the TuPro R&D derived data. From the final MRR front page, the immutable molecular summary report (MSR) is generated (**Figure 3**). Based on this MSR, the clinicians that form the Tumor Board decide whether to follow the pre-TB recommendation or not. The MSR is added to the patient’s clinical record.

## Study Evaluation

In order to generate data that allow for a transparent assessment of the technical performance of each platform and the clinical relevance of the obtained results, aliquots of standardized samples of cultured cells or tumor tissue samples were analyzed by each platform. The resulting data was processed and deposited in the RDMS. The technical performance of each technology has been analyzed based on the parameters below, while the clinical utility will be assessed throughout the study based on utility ratings by the pre-TB and Tumor Board physicians. At the end of the sample and data collection phase, the clinical utility will again be evaluated by means of patient-outcome specific parameters and an extended ESCAT scale (**Supplementary Table 1**).

### Technical assessment

The technical assessment is done at the platform level with the objective of demonstrating that the quality of the data obtained is adequate for the TuPro project. We distinguish three levels of data. We define **Level 1 data** as data with immediate clinical relevance because the improved clinical outcome has been demonstrated in prospective clinical trial(s). These include standard biomarkers predictive of response to an FDA approved direct drug target in a particular indication. **Level 2 data** are defined as data that are likely to have clinical relevance. These include biomarkers for which clinical evidence exists in support of likely clinical benefit, but no or too little data is available on survival endpoints. **Level 3 data** are the remaining part of the acquired data, which likely contains clinically relevant information that may be utilized by data-driven approaches (**Supplementary Figure 8**). An evaluation of technical specifications of each platform for the generation of level 1 and 2 data supporting the clinical assessment (described below) will be performed, including reproducibility, minimum sample requirements, turnaround time, and sample measurement capacity per week. Moreover, the ability to measure known, clinically relevant features exemplified by the measurement of known indication-specific markers or known drug sensitivities will be performed. At a higher level, an assessment of the ability to detect synergies between results from different platforms, exemplified by molecular patterns that mechanistically explain sensitivity or resistance to specific drugs, will be carried out.

### Clinical assessment

The purpose of the clinical assessment of the data is to demonstrate that the additional information provided by TuPro informs and can positively influence clinical decision making. Since an early analysis with a limited number of samples would not have sufficient statistical power and could not be related to clinical parameters such as overall survival (OS) or progression-free survival (PFS), the initial assessment of clinical utility is based on the expert opinion of both a pre-TB and a Tumor Board. Information on the impact of the data provided by the individual TuPro technologies is recorded during the pre-TB and Tumor Board discussions (**Supplementary Table 1, points 1 and 2**). Additionally, at the end of the sample collection and analysis phase, TuPro will be evaluated based on the parameters in Supplementary Table 1 (**Supplementary Table 1, Point 3**). Briefly, overall survival (OS), event-free survival (EFS), the proportion of patients terminating treatment due to toxicity, as well as the quality of life using the FACT-G7 (Yanez et al. 2013) will be assessed. The integrative nature of TuPro allows for the identification of additional markers that could be suggested as novel clinical biomarkers or treatment target candidates, allowing for hypothesis generation and testing within the TuPro framework (**Supplementary Table 1, points 4-6**).

## Exploratory and Integrative Analyses

The integration of clinical information with different types of molecular, cellular and phenotypic data on a tumor sample provides clinically relevant insights that could impact the management of a patient and therefore improve their chances of treatment success (Robinson et al. 2017; Jayawardana et al. 2015). The TuPro study aims to leverage the knowledge accumulated in the research community in order to identify **Level 1** data and to provide strong additional support for **Level 2** data in the enrolled patients through integrative analyses of the datasets collected in this study. This aim will be fulfilled through various efforts that include the development of multi-modal data integration methods for single-cell data. Furthermore, the full TuPro cohort represents a valuable resource to explore the use of **Level 3** data in order to identify putative novel biomarker predictors in cancer (e.g., marker expression, activated pathways, sample composition). Additionally, association studies of data across modalities and detailed analyses of DNA, RNA, protein, and pathway aberrations in cancer will likely lead to a better and more comprehensive understanding of the TuPro tumors’ biology (Lehmann et al. 2015; PCAWG Transcriptome Core Group et al. 2018; Kahles et al. 2018). These analyses can then support clinical trials on these Level 3 data biomarkers for their establishment as Level 1 or 2 data, closing the loop from exploratory science to clinical practice (**Figure 1A**).

## Preliminary results

### Reproducibility and Robustness

Robust and consistent data generation is a crucial prerequisite to ensure the value and reliability of the information provided by TuPro technologies. To demonstrate that reproducible and robust data generation is feasible in TuPro, the single-cell technologies were required to produce consistent and correct cell type fractions, and all TuPro exploratory technologies were tested for their ability to achieve pre-defined quality criteria, across the analyses of one set of triplicates, in the following two settings:

1. **Reproducibility of Cell Type Decomposition:** Artificial cell mixtures with pre-defined proportions (unknown to the participants) of PBMCs and three previously characterized cell lines for melanoma or ovarian carcinoma were prepared and distributed to the TuPro exploratory technologies. All technologies with the capability of differentiating cell types with their marker panels (Pharmacoscopy, single-cell omics, CyTOF, and IMC) determined the proportion of cells corresponding to each cell line. Two technologies that do not aim at identifying cell types still performed the test and inferred cell line fractions based on clustering with data from the analyses of pure cell lines (proteotype analysis) or by grouping cell lines into a single category when markers overlapped (4i DRP). All technologies consistently reported similar cell type fractions (**Supplementary Figure 9A-C**).
2. **Robustness:** To validate robust data generation, every platform produced data based on the artificial sample for melanoma in triplicates. Each technology pre-defined quality criteria thresholds known to be acceptable and achievable by their experimental approach (e.g., coefficient of variation, ARI scores etc.). All technologies were able to attain their target cut-offs (**Supplementary Figure 9D**), which shows that all technologies are able to generate data across a set of triplicates robustly.

### Turnaround Time / Feasibility

The performance metrics of each TuPro technology are shown in **Supplementary Figure 10**. The figure indicates the feasibility of generating data from clinical tissue samples at throughput and timeline that is compatible with clinical decision making, i.e., with a maximum turnaround time from sample reception by the platforms to the finalized analysis of 2 weeks. The TuPro analysis from surgery to pre-TB treatment recommendation is consistently achieved within four weeks.

### Case Study

To date, about 40 patients have been analyzed in the TuPro melanoma study and about 15 patients in the TuPro ovarian study. In the vast majority of cases, the observations reported by the technologies support and complement each other, providing higher confidence in treatment decisions. Furthermore, for most melanoma patients discussed so far in the pre-TB, TuPro exploratory data have provided decision support beyond state-of-the-art and emerging standard clinical diagnostics (histopathology staining and targeted NGS).

This is exemplified by the case of patient TP-M2-USZ-010 with metastatic acral-lentiginous melanoma who progressed on anti-PD-1 therapy and was planned for palliative chemotherapy. A subcutaneous metastasis of this patient was analyzed in the study. Histopathology diagnosed an immune-excluded tumor with PD-1 expression on 80% of T cells and PD-L1 expression on 10% of tumor cells. Targeted NGS revealed low tumor mutational burden (1 Mut/Mb), loss of *NF1* exons 24-29, as well as *ATM, APC*, and *NF2* loss-of-function (LoF) mutations. The sequencing report suggested MEK inhibitors targeting the *NF1* exons 24-29 loss and PARP inhibitors due to the *ATM* LoF mutation. Immunotherapy was not recommended because of the low tumor mutational burden.

Based on these diagnostic data, the pre-TB recommended that due to failed immunotherapy, an immune excluded tumor, low tumor mutational burden, and no apparent targets for small molecule therapy, the patient should undergo palliative chemotherapy as planned before the evaluation by the pre-TB.

The TuPro exploratory technologies CyTOF and Single-cell Genomics & Transcriptomics determined sample composition showing very similar results, both reporting ∼35% tumor cells (in at least 4 clusters), 40% mostly exhausted CD8+ T cells, 12% macrophages, and the remaining fraction representing B lymphocytes and endothelial cells (**Figure 4 A,B**). CyTOF and scRNA-seq showed expression of CTLA-4 and PD-1 on T cells (**Figure 4A, B)**, and IMC could detect a significant interaction of PD-1 positive CD8 T cells with PD-L1 positive tumor cells in four out of six selected regions (p < 0.01, **Figure C**) potentially supporting checkpoint inhibitor treatment. Both CyTOF and scRNA-seq technologies found the melanoma population to be positive for melanocytic phenotype markers such as MITF, MELA, and TYRP1, as well as for Vimentin, indicating a transitional phenotype with mesenchymal features (data not shown). Consistent with the LoF mutation in *APC*, a component of the beta-catenin destruction complex, CyTOF showed a very high beta-catenin signal in the main tumor cluster (**Figure 4A**). While CyTOF could detect pH2AX expression in a small fraction of tumor cells, potentially corresponding to double-strand breaks caused by the reported *ATM* mutation (**Figure 4A**), no increased apoptosis or significant cell killing in the presence of PARP inhibitors could be detected by 4i DRP or pharmacoscopy, respectively. The loss of *NF1* exons 24-29 is supported by loss of NF1 expression in scRNA-seq (not shown); however, whereas MEK inhibitor treatment with Trametinib significantly reduced pERK expression in the 4i DRP analysis (p<0.001, **Figure 4F**) and induced apoptosis, no significant cell death was detected with this treatment by pharmacoscopy (data not shown). Correspondingly, CyTOF analysis showed a low pERK signal in tumor cells (**Figure 4A**), and scRNA-seq showed no overactivation of the MAPK pathway (**Figure 4B**), suggesting that in this case, MAPK pathway signaling might not be the main driver of tumorigenesis.

**Figure 4:**
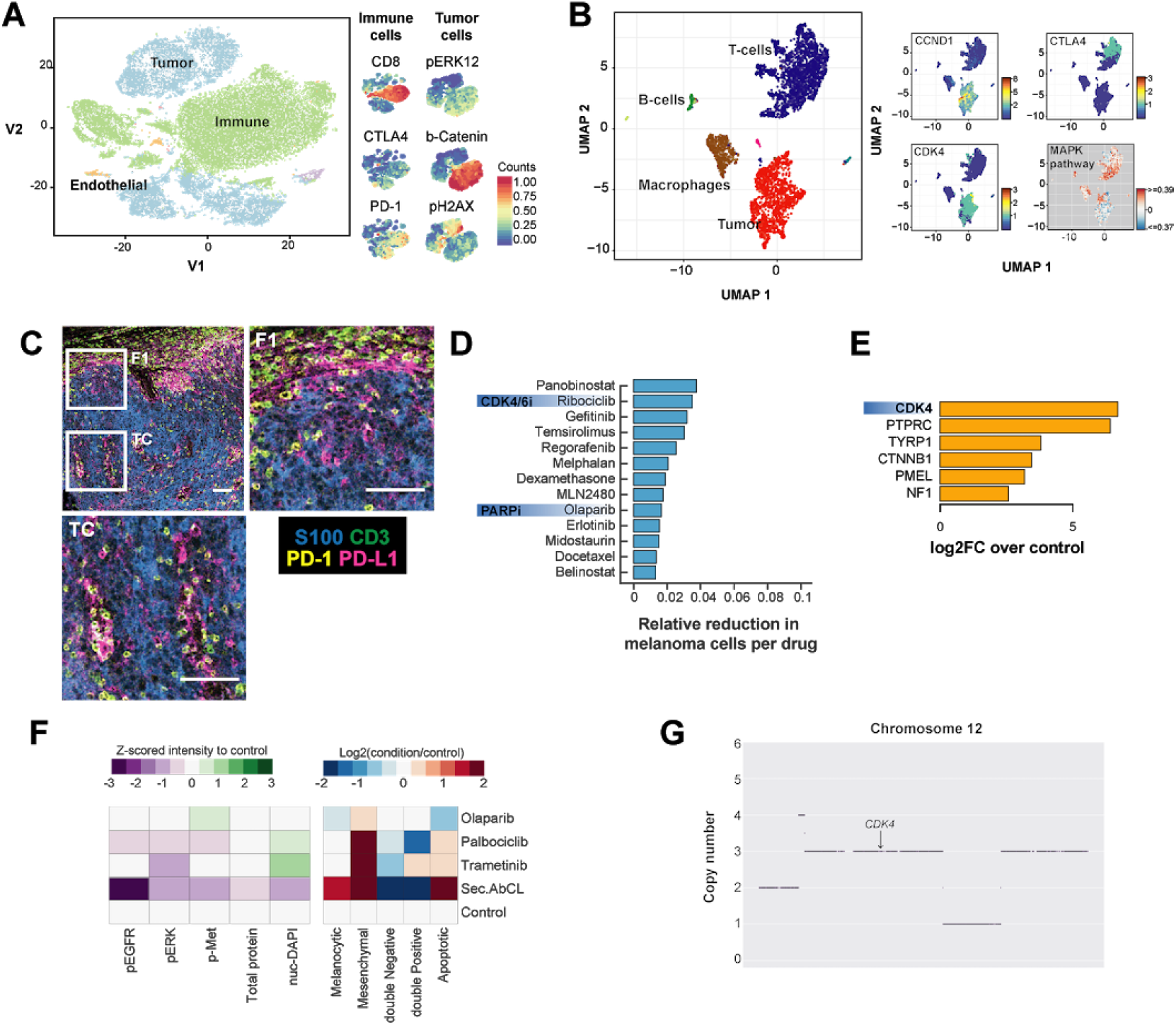
Tumor Profiler technologies provide treatment decision support beyond state-of-the-art diagnostics. Examples from the TuPro analysis of melanoma patient TP-M2-USZ-010. A) and B) CyTOF TSNE plots and scRNA-seq UMAP plots show consistent tumor composition (40% tumor, 40% T cells, 12% macrophages) as well as evidence for CTLA-4 expression and CDK4/6 pathway activation. C) IMC indicates an interaction of PD-L1+ tumor cells with PD-1+ T cells. Interaction was significant by permutation testing in four of six regions of interest, i.e. in these regions the interaction score was higher compared to random shuffling (p<0.01). D) Pharmacoscopy analysis indicates CDK4/6 inhibitor as one of the top hits, however, for this sample, none of the drugs reached significance (p>0.05, one-tailed t test) in inducing cell death compared to DMSO control. E) Proteotype analysis confirms a higher abundance of CDK4 compared to control. Detection of NF1 peptides might be due to high immune content, as indicated by overrepresentation of PTPRC (CD45) peptides. F) 4i DRP analysis indicates increased apoptosis and downregulation of pERK (p<0.001), pMET (p<0.001), and pEGFR (p<0.0001) upon Palbociclib (CDK4/6i) treatment. G) scDNA-seq shows that CDK4 is located in a large genomic region with a low-level copy number gain (CN=3).

Interestingly, proteotyping observed high CDK4 protein expression (**Figure 4E)** in the tumor; and correspondingly scRNA showed consistent expression of CDK4 mRNA in the tumor clusters (**Figure 4B)**. Moreover, scDNA-seq detected a low-level copy number gain (CN=3) in a large chromosomal region containing *CDK4* (**Figure 4G**). This alteration was not reported by bulk targeted NGS as it is below the panel’s reporting threshold for clinically relevant CNVs (Frampton et al. 2013). Importantly, and consistently with high CDK4 expression, both drug profiling technologies detected sensitivity to the CDK4/6 inhibitors *Ribociclib* or *Palbociclib*, i.e. increased apoptosis and significant oncogenic pathway downregulation (p<0.001) as well as increased melanoma cell death in 4i DRP and pharmacoscopy, respectively (**Figure 4F, D**).

Taken together, the described genetic alterations are known in melanoma but are among the less frequent melanoma drivers with an average frequency of <10% (Cancer Genome Atlas Network 2015). Genetic loss of function alterations in *NF1* are activators of the MAPK and PI3K/AKT pathways (Cirenajwis et al. 2017), which is in line with the observed low sensitivity to single MEK inhibitor treatment. Genetic alterations in *ATM* are frequently associated with impaired homologous recombination qualifying for PARP inhibitor treatment (Michels et al. 2014), but a role for ATM has also been described in the negative regulation of Cyclins (Beishline and Azizkhan-Clifford 2014). As such, the *ATM* LoF mutation identified in the present case might have contributed to the observed increased Cyclin D1 levels (Fig. 4B), potentially resulting in an activation of the CDK4/6 pathway and CDK4/6 inhibitor sensitivity. In addition to targeting cell proliferation, CDK4/6 inhibitors may increase tumor immunogenicity (Goel et al. 2017). This effect is of particular interest in an immune excluded tumor as the one described. In view of all data available, including supportive exploratory data for immunotherapy, the pre-TB, therefore, made the treatment recommendation of an off-label CDK4/6 inhibitor in combination with approved anti-PD-1 plus anti-CTLA-4 therapies. The Tumor Board followed the treatment recommendations of TuPro’s pre-TB in principle. However, due to clinical deterioration, active anti-tumor treatment was not an option, and the patient received palliative chemotherapy.

This case study exemplifies how data generated by the individual TuPro technologies can be utilized in an integrative manner and, importantly, can provide therapeutic decision support that complements emerging standard diagnostic techniques. In this case, the TuPro exploratory technologies not only provided additional evidence on the functional significance of the genetic variants detected by Targeted NGS but have also contributed further information that has allowed treatment recommendations that were not considered before based solely on genetic data. The increment of the explorative TuPro technologies might be particularly valuable for cases where the effect of oncogenic drivers is weak, has pleiotropic effects, where it generally remains challenging to single out the primary driving pathway, or on pre-treated tumors where the tumor-maintenance mechanisms are harder to identify.

## Discussion and Outlook

Treatment decisions based on histopathological analyses and targeted NGS results are quickly becoming standards in tumor boards worldwide. However, our understanding of the complex interactions that comprise the tumor microenvironment and its response to targeted or immunotherapies is still in its infancy. For this reason, the TuPro consortium has built a state-of-the-art profiling platform that captures cellular, molecular, spatial, and functional information from three tumor types and generates molecular research and summary reports for a tumor board presentation. Not only have these data enhanced the results of the routine diagnostics that are typically used to select treatments, but they will also provide a powerful database of single-cell, high-dimensional, and functional information for exploratory analyses including biomarker discovery. In the context of an observational, technology demonstration clinical study, TuPro reports integrating these multi-omic analyses are presented in pre-TBs in which hypothetical treatment decisions will be recorded for all patients. An integrated analysis will be carried out during the course of the study in one indication becomes available. The TuPro team will compare results from emerging clinical standard technologies (i.e., histopathology and targeted NGS) on their own to the same results considered along with data from TuPro technologies. The comparative analyses of multi-level datasets between patients who responded and those who did not respond to therapy may allow the identification of potential mechanisms of resistance to standard treatment and may thus allow for the identification of potential novel therapeutic targets, as illustrated in the case study. Although the cohort will still be relatively small at this point of the study, it will allow for the generation of hypotheses that can be tested in subsequent patient samples as well as in publically available datasets or other reference cohorts.

In order to capture the influence of TuPro results in clinical decision making, we plan to adapt the ESCAT scale (Mateo et al. 2018) routinely used to rank genomic alterations as targets of precision medicine to take into account other types of molecular, cellular, and functional data as measured by TuPro emerging technologies. The relevance of the observations gathered throughout TuPro in treatment prediction will directly inform the creation of this adapted clinical actionability scale.

As our knowledge of tumor biology advances and our ability to specifically target its features becomes more sophisticated, it is imperative that we develop better tools to more accurately describe and model the complex tumor microenvironment at various phenomenological levels. However, if we want to achieve an impact on clinical practice and ideally improve the therapeutic options for cancer patients, these data need to be generated, interpreted, and summarized in fast-paced clinical environments with different ethical, regulatory, and temporal constraints than are present in academic research labs. For this reason, the TuPro consortium was designed to meet the demands of clinical practice and to produce rich, high-dimensional datasets for in-depth exploratory analyses. Not only will this approach have the chance to change the future of cancer patient management by providing more individualized therapies, but it will also facilitate the identification of novel prognostic or predictive biomarkers and potential new drug targets. Although melanoma patients, in particular, have substantially benefited from targeted and immunotherapies over the last several years, even greater improvements in patient survival across many cancer types will surely result from the implementation and integration of complementary, cutting-edge technologies into clinical practice.

## Data Availability

The manuscript details a prospetive outlook for a study that is currently underway. However, the data will be made available upon study completion and publication.

## Acknowledgements

This project is jointly funded by a public-private partnership involving Roche, ETH Zurich, University of Zurich, University Hospital Zurich and University Hospital Basel. We would like to acknowledge the support of the Biobank team from the Department of Dermatology, and The Biobank and Tumor Genomic Profiling team of the Department of Pathology and Molecular Pathology at The University Hospital Zurich. Moreover, we acknowledge the support of the Clinical Trials Center (CTC) Zurich and the Clinical Trials Unit (CTU) Basel. We gratefully acknowledge the excellent project management support by the following individuals: Andreas Strauss, Thomas Solbach, Lina Reypens, and Vanessa Mareike Johnen.

## Author contributions

### Conceptualization of study

Rudolf Aebersold, Niko Beerenwinkel, Christian Beisel, Bernd Bodenmiller, Holger Moch, Mitchell P. Levesque, Lucas Pelkmans, Gunnar Rätsch, Berend Snijder, Markus Tolnay, Bernd Wollscheid, Gregor Zünd, Gabriela Senti, Detlef Günther, Christoph Hock, Beatrice Beck-Schimmer, Andreas Wicki, Reinhard Dummer, Werner Kübler, Sebastian Lugert, Maya D’Costa, Marina Bacac, Peer-Olof Attinger, Severin Schwan, Tim M. Jaeger, Christian Rommel, Gerd Mass

### Preliminary results

- Robustness/Replication: Jack Kuipers, Mustafa A. Tuncel, Franziska Singer, Anne Bertolini, Sandra Goetze, Julien Mena, Emanuela Milani, Rebekka Wegmann, Ruben Casanova, Nunu Mchedlishvili, Gabriele Gut, Jacobo Sarabia del Castillo, René holtackers, Katja Eschbach, Gunnar Rätsch, Kjong Lehmann, Ximena Bonilla, Joanna Ficek, Andrea Jacobs, Stefanie Engler, Sujana Sivapatham, Stéphane Chevrier
- Case study: Anja Irmisch, Johanna Ziegler, Ximena Bonilla, Nora Toussaint, Gunnar Rätsch, Mitchell P. Levesque, Viktor H. Koelzer, Bettina Sobottka, Reinhard Dummer
- Data analysis: scGenomics: Jack Kuipers, Ulrike Menzel, Michael Prummer, Franziska Singer, Mustafa A. Tuncel, Christian Beisel, Niko Beerenwinkel; Proteotype analysis: Emanuela Milani, Sandra Goetze, Bernd Wollscheid; Pharmacoscopy: Rebekka Wegmann, Julien Mena, Berend Snijder; 4i DRP: Cinzia Esposito, Gabriele Gut, Jacobo Sarabia del Castillo, René Holtackers, Lucas Pelkmans; IMC: Ruben Casanova, Shuhan Xu, Bernd Bodenmiller; CyTOF: Stéphane Chevrier, Shuhan Xu, Bernd Bodenmiller; Integrative and exploratory analyses: Kjong-Van Lehmann, Ximena Bonilla, Joanna Ficek, Gunnar Rätsch; Clinical science: Anja Irmisch, Francis Jacob, Mitchell P. Levesque, Reinhard Dummer, Andreas Wicki, Michael Weller; cfDNA: Salvatore Piscuoglio, Charlotte KY Ng, and Jermann Philip Martin; Data/IT: Daniel J. Stekhoven, Nora C. Toussaint, Gunnar Rätsch; Digital Pathology: Viktor Hendrik Koelzer, Bettina Sobottka, Daniel Baumhoer, Stefan Nicolet, Anja Frei, Holger Moch

### Manuscript writing

Anja Irmisch, Ximena Bonilla, Stephane Chevrier, Kjong-Van Lehmann, Franzisca Singer, Nora C. Toussaint, Cinzia Esposito, Julien Mena, Emanuela Milani, Ruben Casanova, Daniel Stekhoven, Rebekka Wegmann, Bettina Sobottka, Sandra Götze, Jack Kuipers, Jacobo Sarabia del Castillo, Ulrike Menzel, Viktor Hendrik Koelzer, Rudolf Aebersold, Mitchell P. Levesque, Gunnar Rätsch

### Writing team coordination

Gunnar Rätsch, Mitchell P. Levesque, Ximena Bonilla, Anja Irmisch

### Manuscript Approval

All authors read and approved the manuscript.

## Supplementary Material

### Supplementary Figures

**Supplementary Figure 1:**
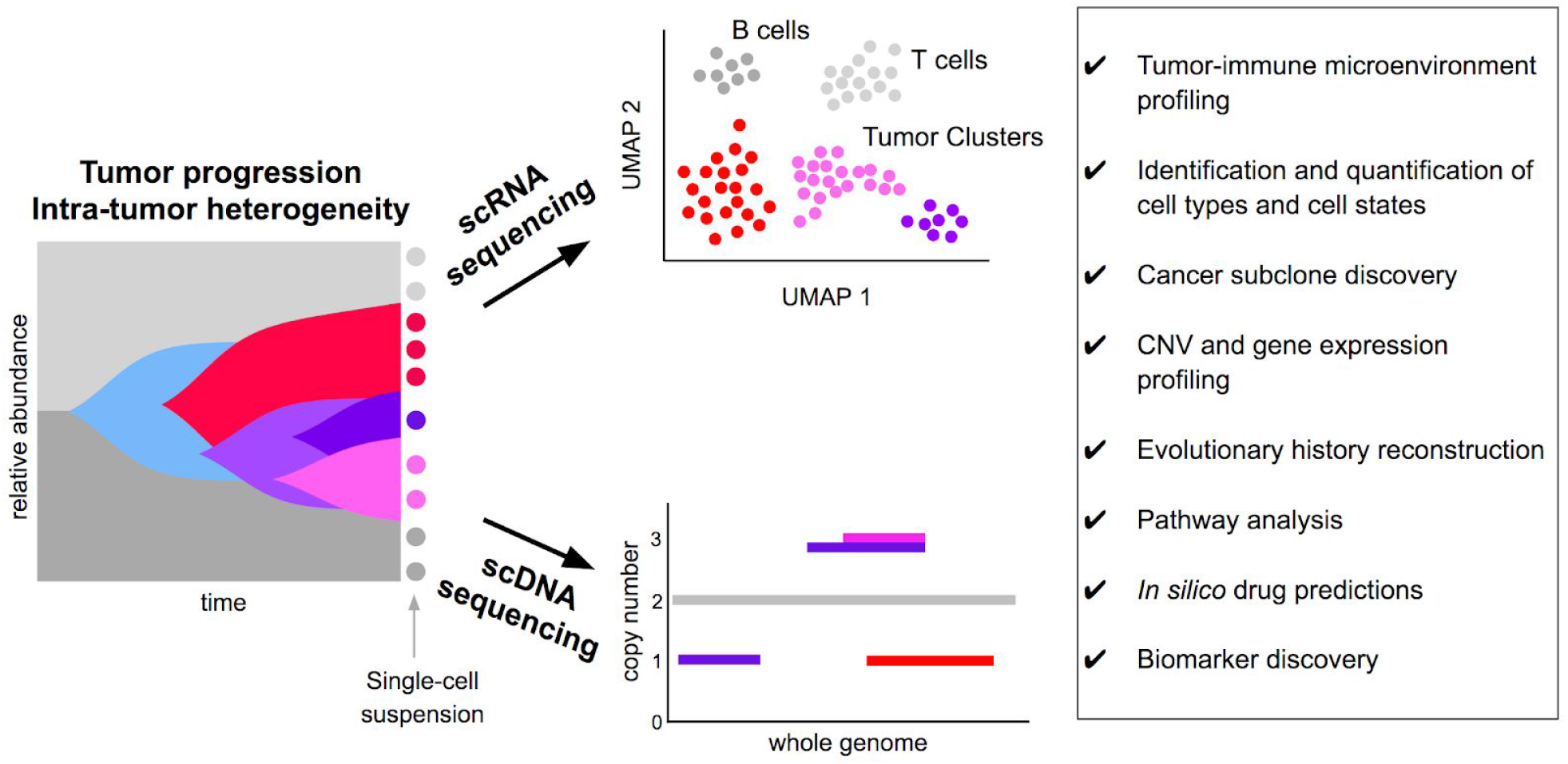
Schematic representation of the Single-cell Genomics & Transcriptomics node. Based on the single-cell suspension of the sample, droplet-based single-cell sequencing of the messenger RNA and genomic DNA is performed, targeting approximately 4000 cells for scRNA-seq and 250 cells for scDNA-seq, respectively. For scRNA-seq, the resulting gene expression counts per cell are quality controlled and normalized, to allow a cell type classification per cell and an unsupervised clustering to infer the sample’s tumor-immune composition and tumor heterogeneity. For scDNA-seq, reads are quality controlled, normalized and binned across the whole genome to allow for copy number calling per cell and phylogenetic reconstruction of the tumor architecture. For details on the readouts, see box.

**Supplementary Figure 2:**
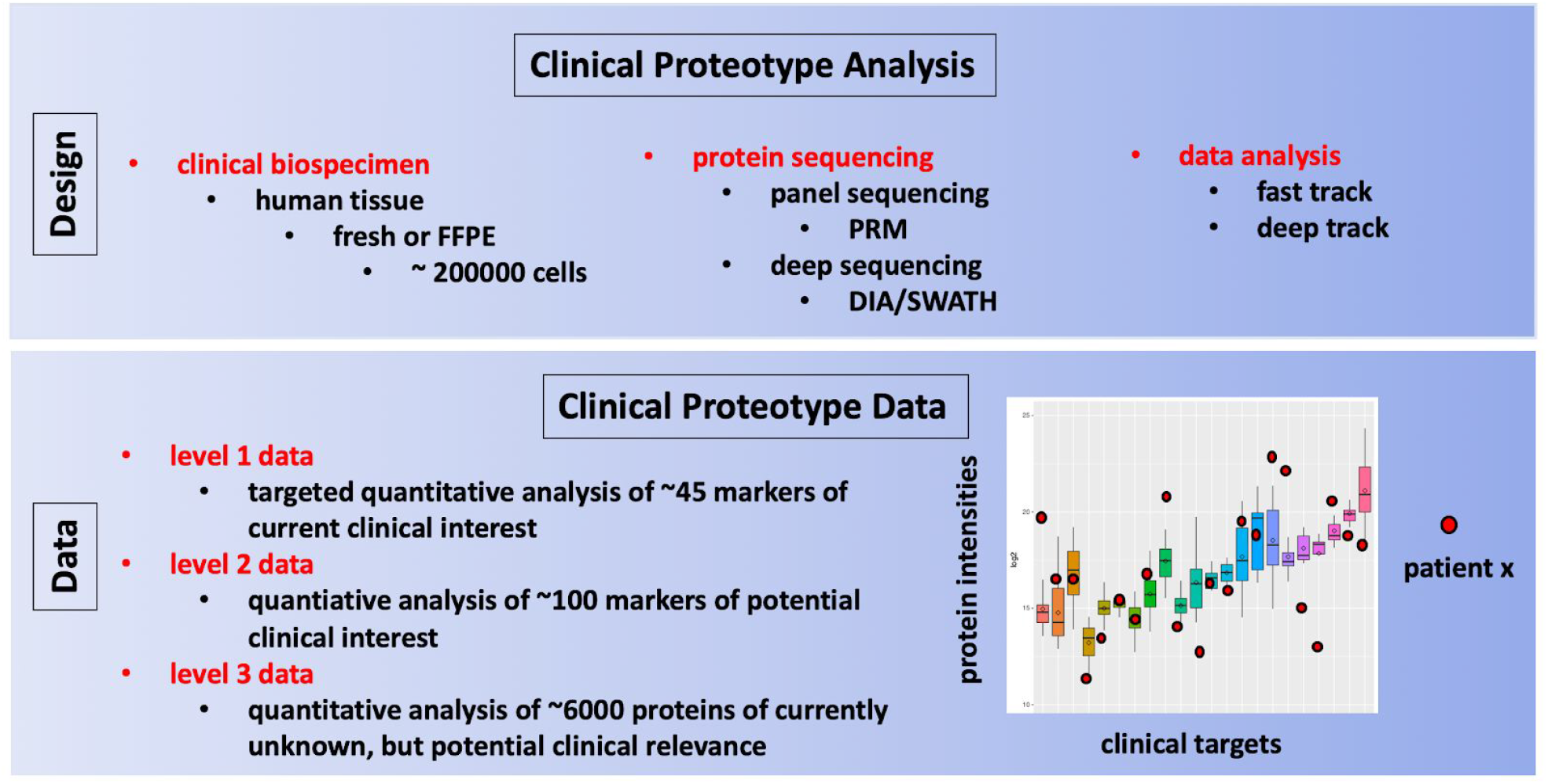
Schematic representation of the Proteotype analysis platform workflow. FFPE tissue samples and/or dissociated cells from tissues are digested into peptides and combined with reference peptides for subsequent analysis by targeted (PRM) and untargeted (DIA/SWATH) quantitative mass spectrometry workflows. Spectral data obtained from the mass spectrometer is processed via fast track (identity and quantity of proteins) and deep track (post-translational modifications; peptidoforms, functional signaling network interrogation) bioinformatic data analysis strategies. The resulting data indicates relative quantitative protein abundance values for the abundance of 45 protein markers of current clinical interest (level 1 data; drug targets), 100 protein markers of potential clinical interest (level 2 data; discussed in the literature, but not in used in clinical practice yet) and 6000 proteins present in the tissue sample of currently unknown, but potential clinical relevance (level 3 data). The data is visualized per patient and per protein in relation to a reference protein abundance range established across a control group. Alone and integrated with data from the other TuPro nodes proteotype profiles generated across the observational clinical trial will be evaluated for their informational value in predicting drug response.

**Supplementary Figure 3:**
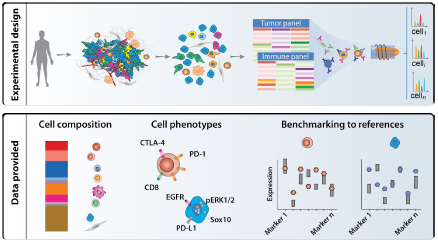
Schematic representation of the CyTOF platform workflow. Dissociated cells are stained together with reference cells with two metal-tagged antibody panels to characterize the tumor and immune cells, respectively. Upon staining, the cells are analyzed by mass spectrometry (upper part). Single-cell data are analyzed to provide the tissue composition, and the signature of each cell type identified, which is then compared to the reference cells included in each individual experiment (lower part).

**Supplementary Figure 4:**
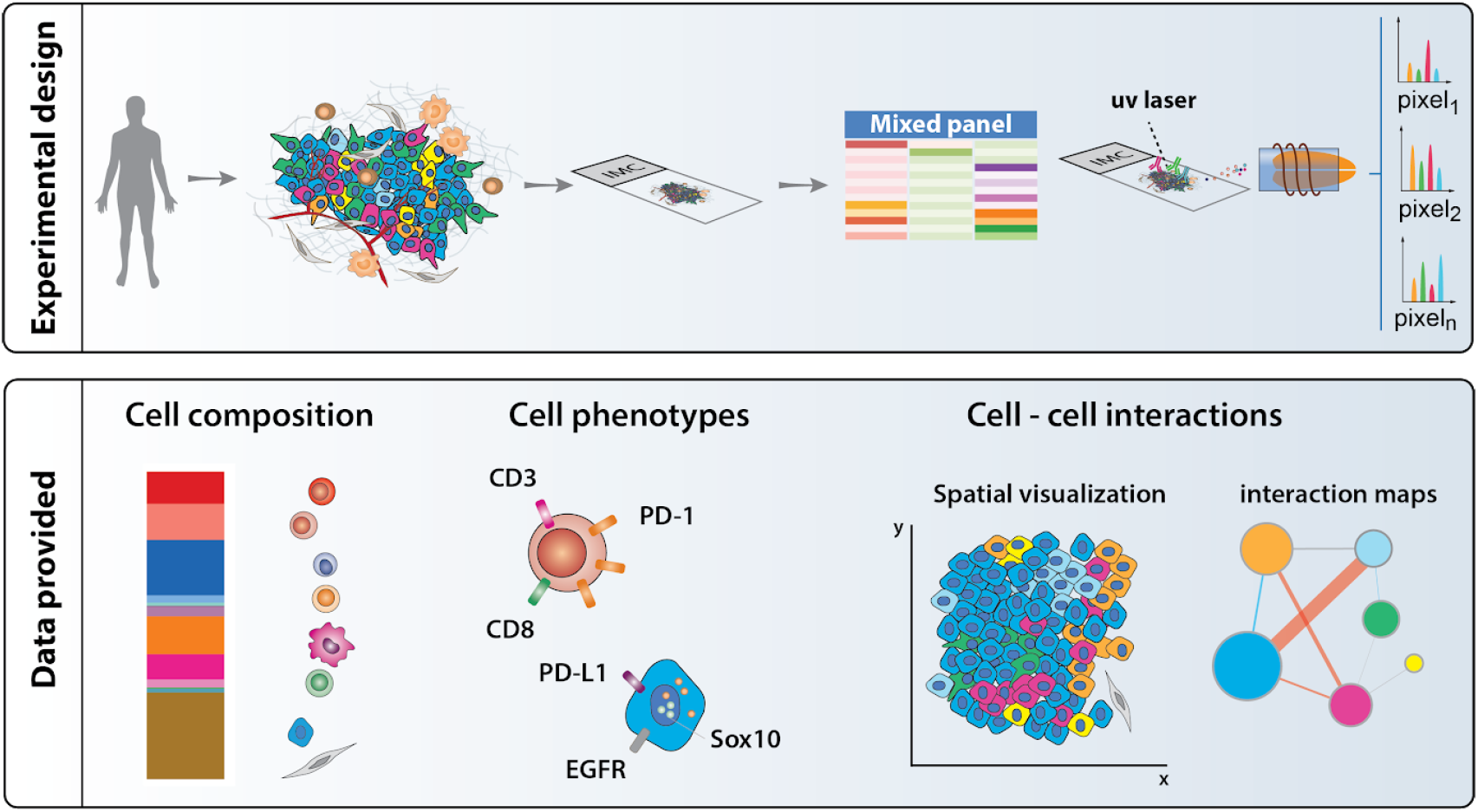
Schematic representation of the Imaging Mass Cytometry (IMC) platform workflow. Formalin-Fixed Paraffin-Embedded (FFPE) tissue sections are stained with a panel of metal-tagged antibodies to characterize both immune and tumor cells. Upon staining, the tissue is ablated with a laser and analyzed by mass spectrometry (upper part). Single-cell data are analyzed to provide the tissue composition, signature of each cell type and quantification of interactions between cell types (lower part).

**Supplementary Figure 5:**
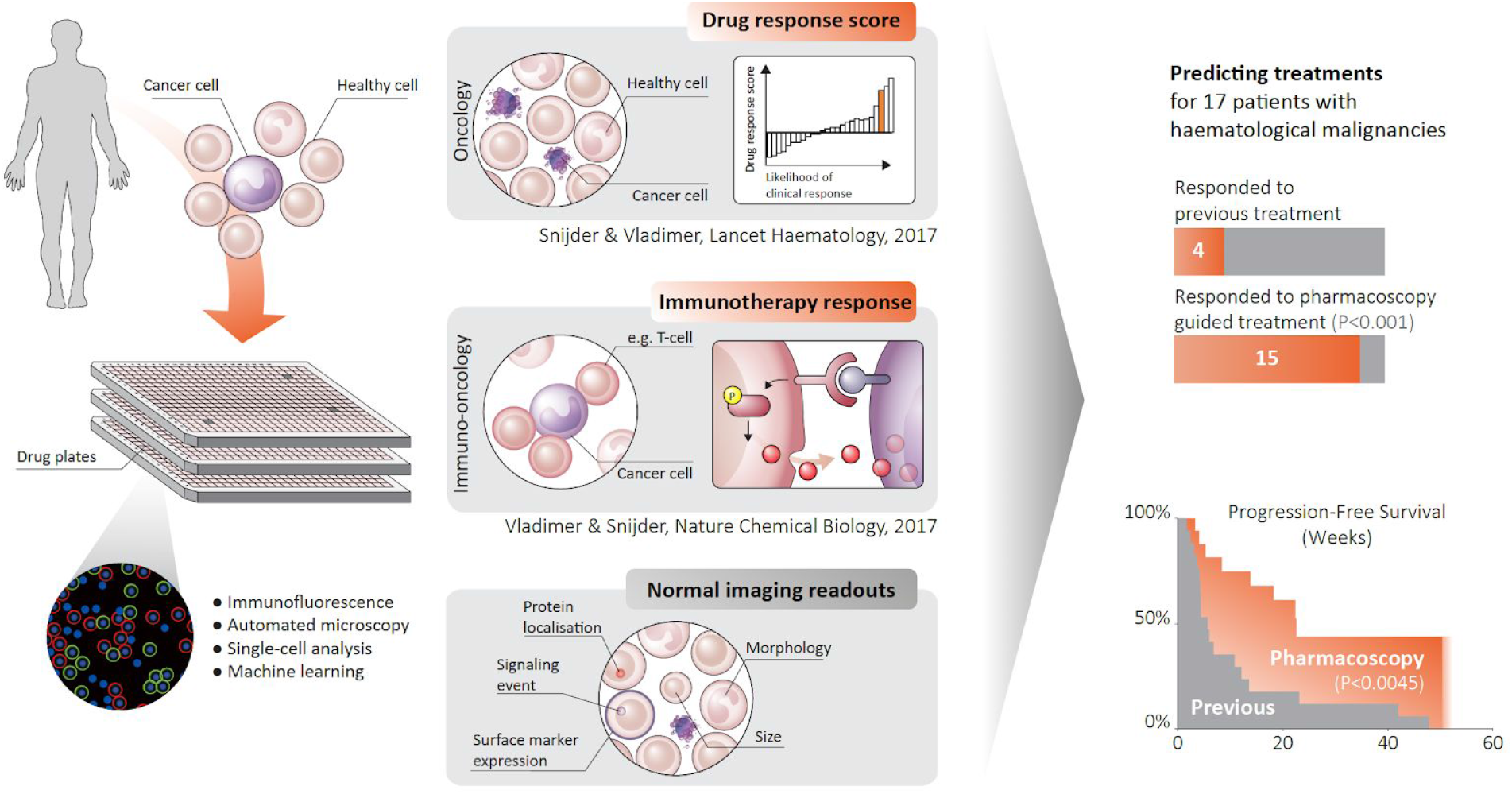
Schematic representation of the pharmacoscopy workflow. A single cell suspension of patient-derived tumor material is transferred to a 384-well plate containing many different drugs and drug combinations with multiple concentrations and replicates per compound. After incubation for 24 hours, the cells are fixed and stained with fluorescent antibodies against tumor markers or counterstains against healthy cells. The full plate is then image using high-throughput automated microscopy. Computational image analysis is used to identify different cell types and marker expression, as well as spatial features such as cell-cell interactions. Furthermore, drug responses are assessed on a single-cell level. To identify the most promising treatments, changes in the fraction of cancer cells upon treatment are assessed. Drugs with on-target effect that specifically kill the cancer cells, but not the healthy cells, receive a high score, whereas ineffective drugs are scored as 0 and drugs that are more toxic to healthy cells than to cancer cells result in a negative score. Using such drug ranking has previously been shown to improve response rates and progression free survival in a cohort of 17 patients with hematological malignancies (Snijder & Vladimer, The Lancet. Haematology, 2017).

**Supplementary Figure 6:**
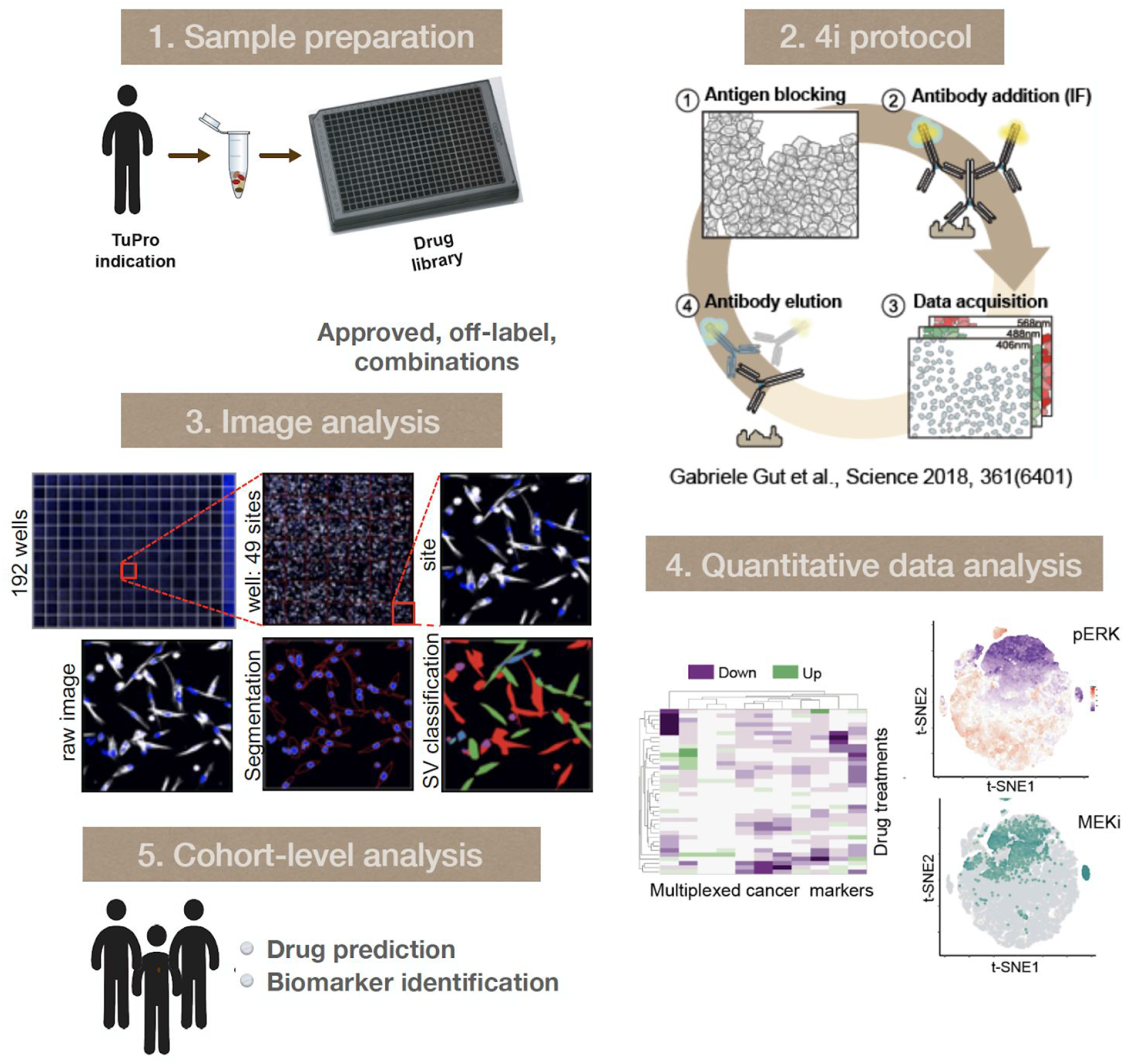
Schematic of the 4i Drug Response Profiling (4i DRP) platform. 1) The tumor cells are plated up in a 384 well-plate format and let to attach overnight before drug treatment. Drug library is tailored to each cancer indication and includes approved, off-label and drug combinations. 2) 4i protocol allows for multiplexed antibody probing against key markers of cancer proliferation. 3) Computational image analysis with multiscale cell detection, object segmentation and identification of phenotypic classes of interest. 4) Quantitative multivariate analysis of single-cell features. Aggregate (left panel) and single-cell (right panel) response of cancer proliferation markers versus drug treatment are generated. 5) On a larger cohort of patients, comparative analysis is carried out to identify diagnostic biomarkers and predict drug response.

**Supplementary Figure 7:**
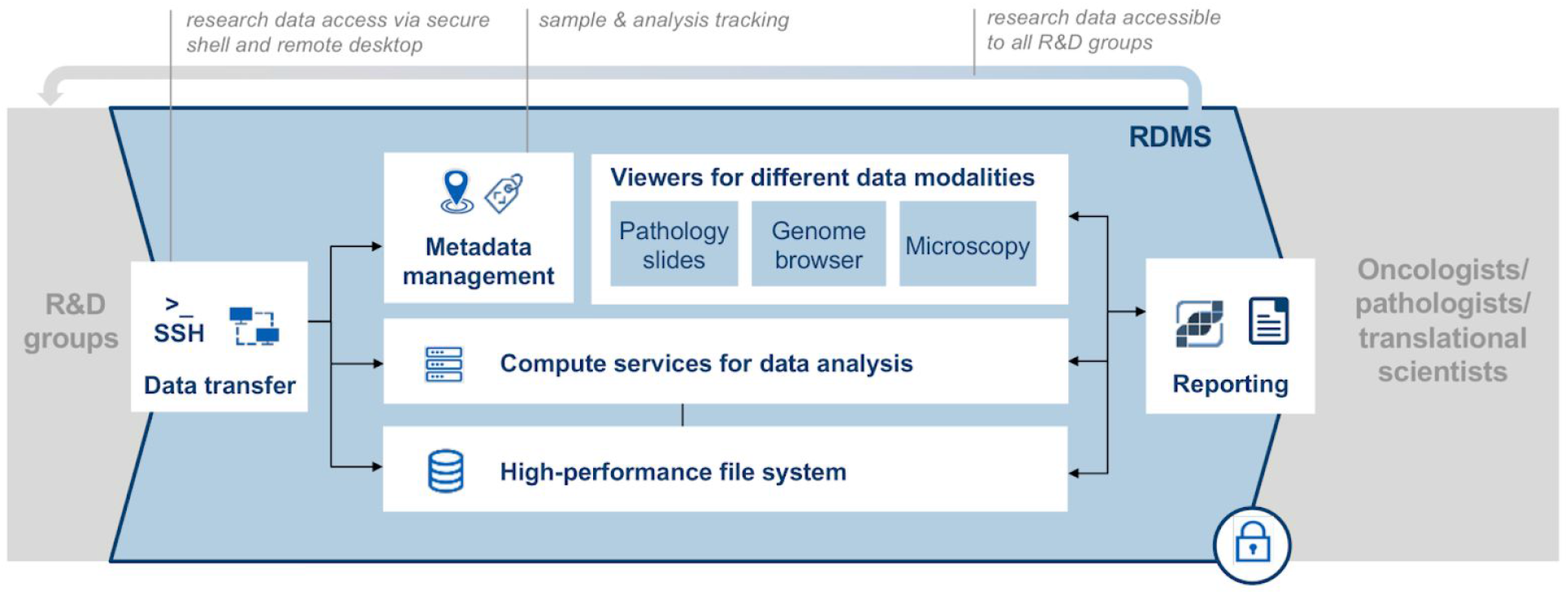
Overview of the TuPro Research Data Management System. The Research Data Management System is a system of software and hardware that securely stores, manages, and allows access to the data generated in the TuPro project. It comprises high-performance file system and compute resources, data viewers (including report viewer), metadata management for sample, and analysis tracking.

**Supplementary Figure 8:**
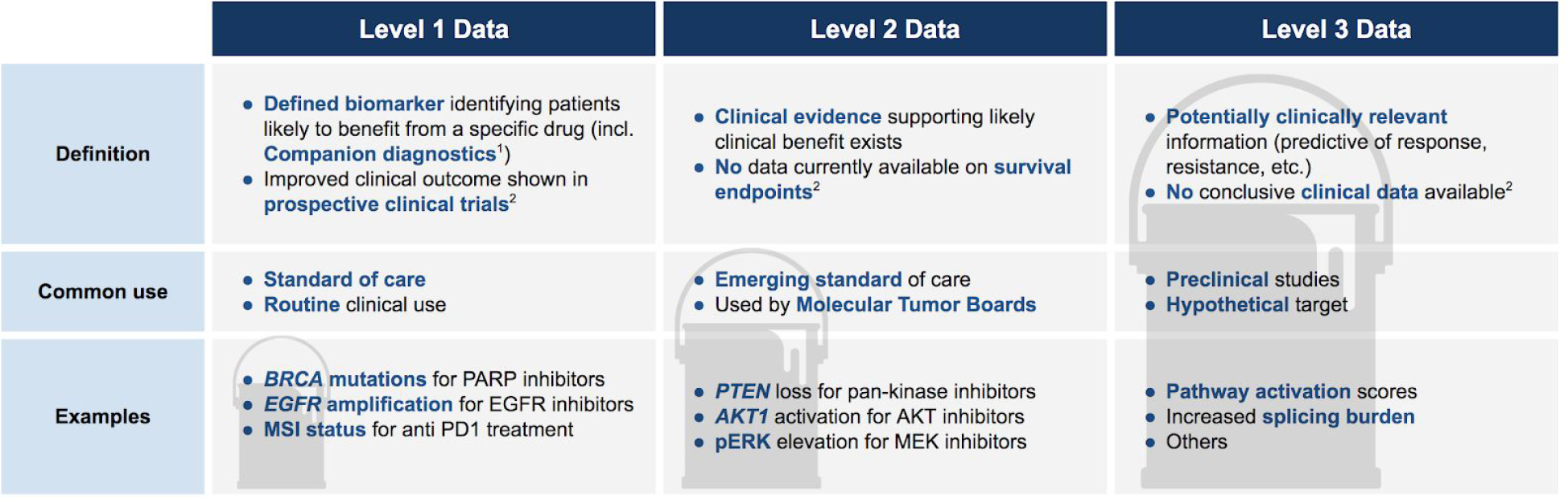
Levels of clinical utility for different types of molecular data. We define three levels of data based on the level of evidence available on their utility (Definition section). The Common use section defines the current level of usage in the medical diagnostics community, while the last section provides representative examples for each one of the three data categories. The bucket image in the background represents the amount of data available for each kind. We expect that the amount of level 1 data is small and has been already characterized to a certain extent. Level 2 data requires additional data integration and analysis approaches in specific contexts and is currently being mined by researchers and clinicians, while level 3 data is mostly unexplored at the clinical utility level and requires large studies and somewhat more complex integration approaches to yield clinical-level results. MSI: microsatellite instability ^1^ https://www.fda.gov/regulatory-information/search-fda-guidance-documents/vitro-companion-diagnostic-devices ^2^ Mateo et al., 2018

**Supplementary Figure 9:**
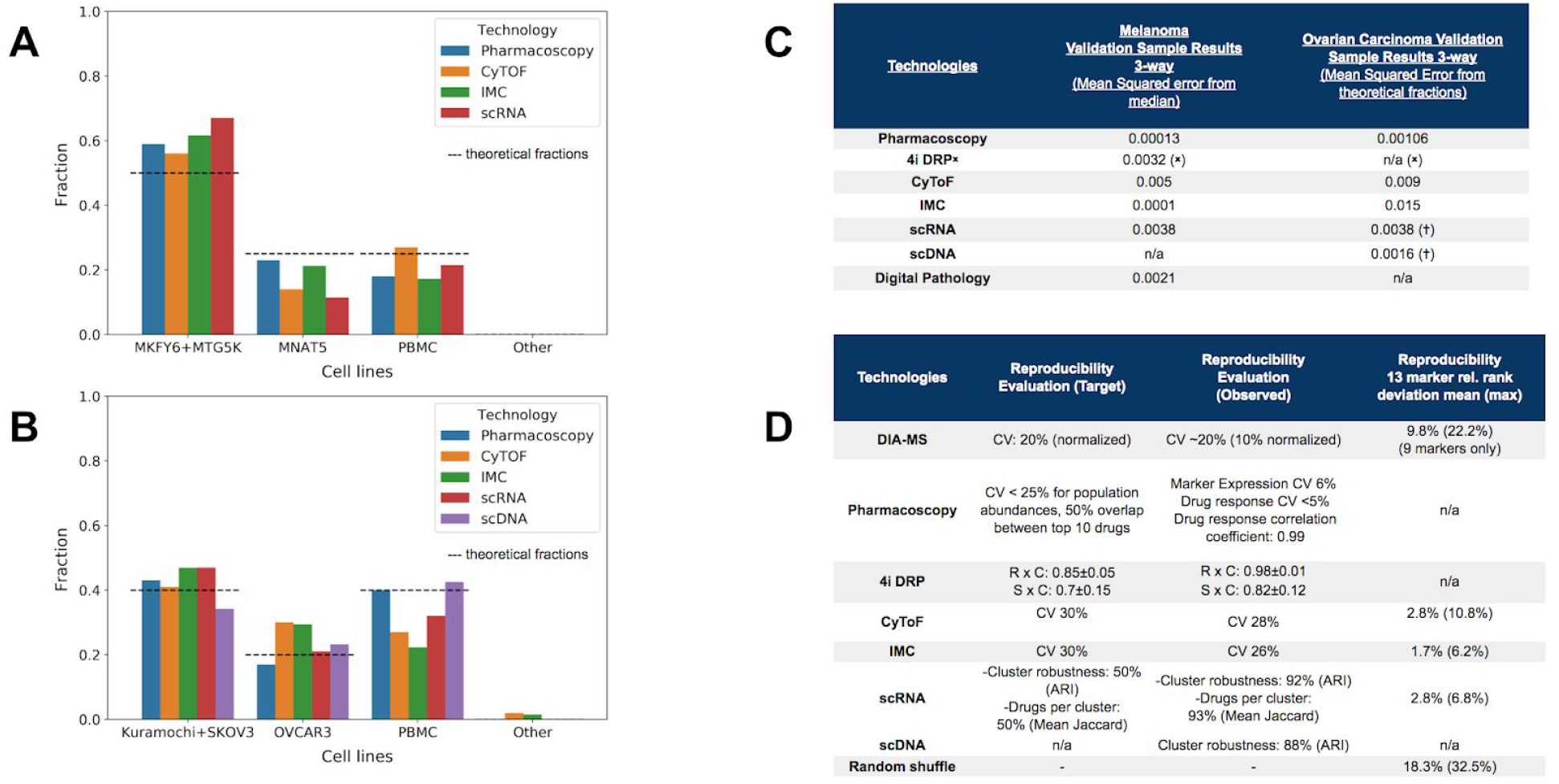
Robustness and Reproducibility. **A)** Subclonal composition variability in Melanoma. **B)** Subclonal composition variability in Ovarian Carcinoma. **C)** Table summarizing the results of the cell-type decomposition test for melanoma and ovarian carcinoma. ᕁNo PBMC marker available, washing steps removing most PBMCs during sample preparation lead to altered expected proportions, †cell-line assignment performed post-hoc based on identified cell fractions (no sequencing of pure cell-lines was done by these technologies and thus the expected expression or copy number profile was not known). Proteotype was not included in this test because the identification of subclonal population of cells is not easy to infer reliably from a bulk technology. **D)** Each node capable of discriminating among cell types in a cell mix, analysed a set of triplicates for reproducibility (here shown for marker expression).The two left columns show the reproducibility target, defined as the current state of the art for each technology, and the observed numbers. A rank based assessment of marker expression reproducibility on a cross-node overlapping set of 13 markers give a sense of trade-offs between resolution and accuracy. Proteotyping has high resolution (5,000 proteins) but higher relative rank deviation while CyTOF and IMC each have considerable lower resolution (2×30 markers) but on single cell resolution with a lower relative rank deviation. scRNA identifies gene expression across 20,000 genes on single cell resolution with lower relative rank deviation.

**Supplementary Figure 10:**
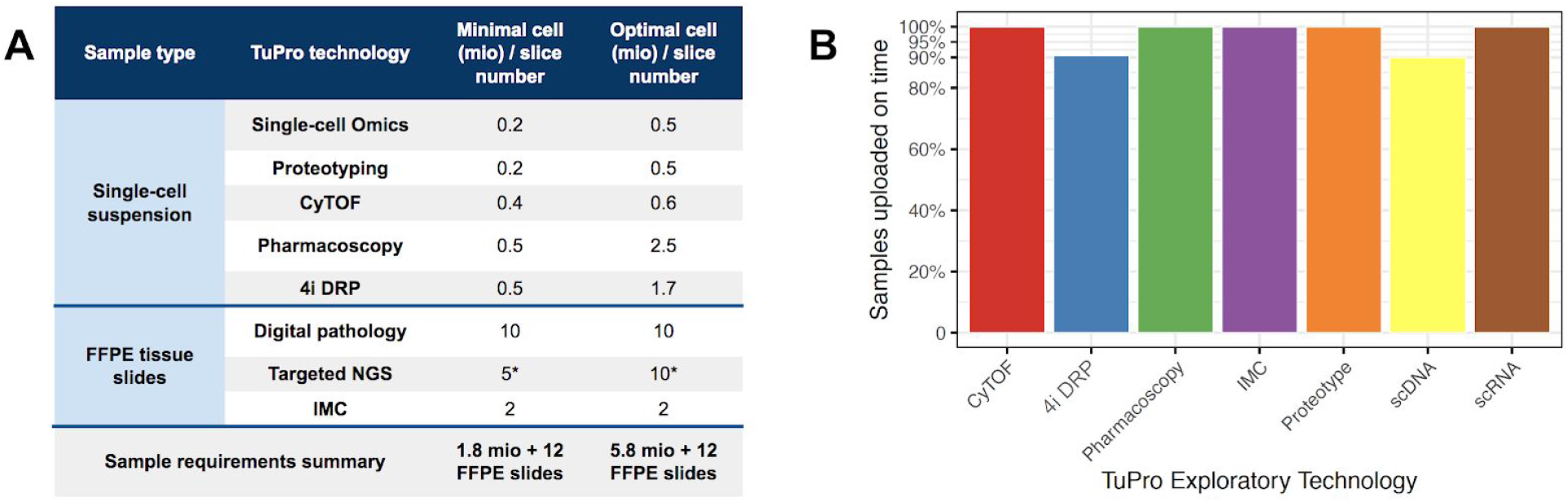
Performance metrics of the TuPro and emerging technology platforms. **A**) The platform name, as well as the type of material required (single cell suspension or formalin-fixed, paraffin embedded -FFPE tissue slides) is noted on the left. The subsequent columns display minimum and optimal amount of material needed to deliver quality results per technology. At the bottom of the table is possible to see a summary of the total cell number and FFPE tissue slides needed for all technologies to provide results. *Targeted NGS requires 5-25 mm2 of 5-10 unstained slides (4-5 u thickness) to extract at least 30 ng and optimally 50 ng of DNA **B**) Percentage of samples uploaded on time to be included in the molecular report from the total number of samples analysed that produced an interpretable result for the melanoma indication.

### Supplementary Tables

**Supplementary Table 1:**
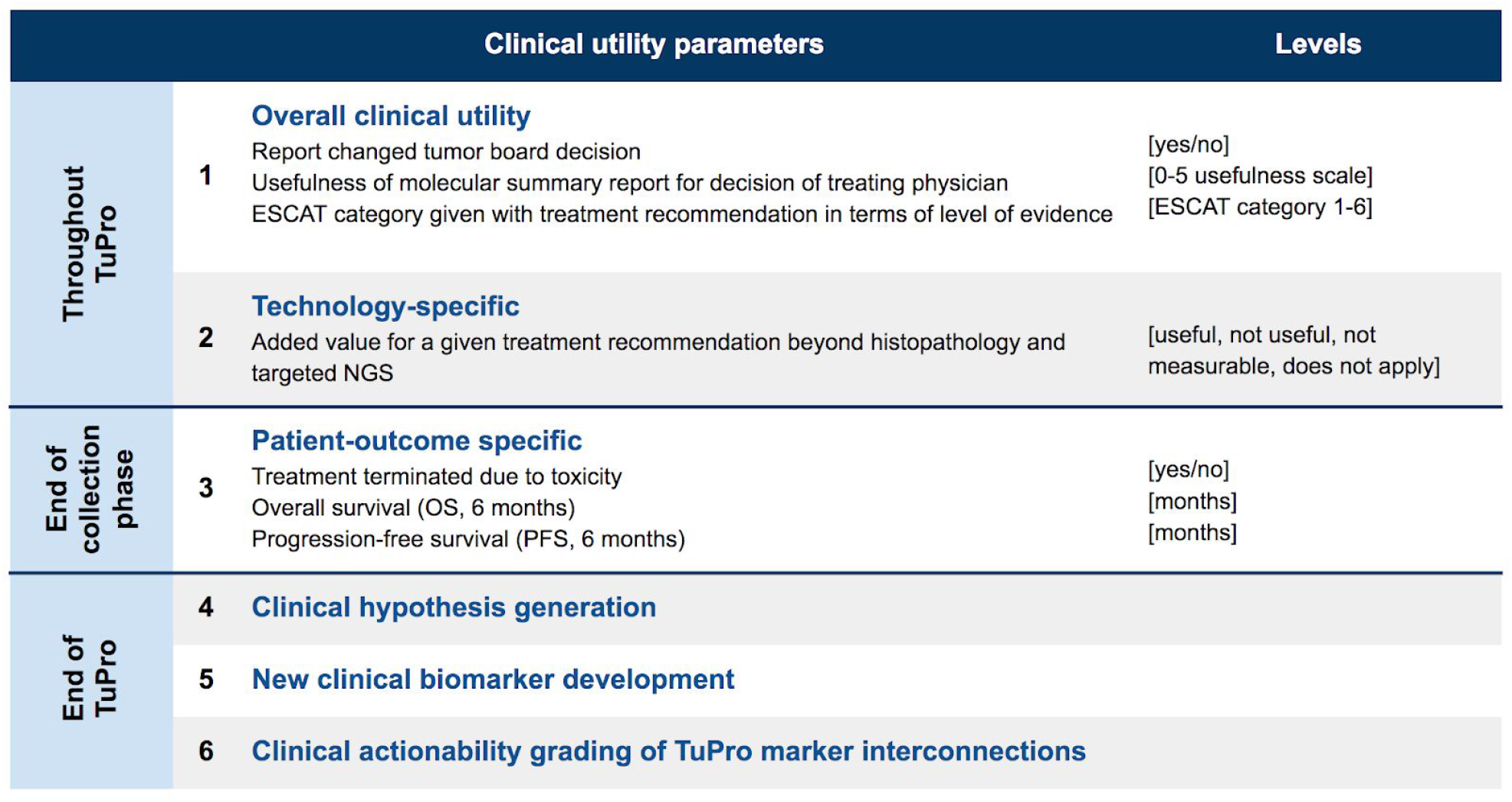
Clinical utility is assessed in 6 different dimensions (points 1-6). The recorded values are listed in the “Levels” column. The two initial clinical utility parameters points represent information that is collected and assessed throughout the study. Point 3 information is analysed at the end of the collection and analysis phase, and points 4-6 are investigated once enough samples are collected, most likely at the end of the study, once all the information has been integrated.

| Clinical utility parameters |  | Levels |
| --- | --- | --- |
| Throughout<br>TuPro | **Overall clinical utility**<br>1 Report changed tumor board decision<br>Usefulness of molecular summary report for decision of treating physician<br>ESCAT category given with treatment recommendation in terms of level of evidence | [yes/no]<br>[0-5 usefulness scale]<br>[ESCAT category 1-6] |
|  | **Technology-specific**<br>2 Added value for a given treatment recommendation beyond histopathology and targeted NGS | [useful, not useful, not measurable, does not apply] |
| End of<br>collection<br>phase | **Patient-outcome specific**<br>3 Treatment terminated due to toxicity<br>Overall survival (OS, 6 months)<br>Progression-free survival (PFS, 6 months) | [yes/no]<br>[months]<br>[months] |
| End of<br>TuPro | 4 **Clinical hypothesis generation** |  |
|  | 5 **New clinical biomarker development** |  |
|  | 6 **Clinical actionability grading of TuPro marker interconnections** |  |

### Supplementary Note 1

The identification and molecular analysis of circulating tumor DNA (ctDNA) in blood is a useful minimally-invasive method to monitor tumors and disease progression (Stewart et al. 2018). Plasma samples from individuals participating in TuPro are profiled with indication-specific Ion AmpliSeq HD custom gene panels (Iontorret, n.d.). The melanoma panel spans 11 genes (97.4% of the CDS covered on average, primary genes such as TP53 and B2M are covered at 100%) and 123 regions of interest (most regions covered at 100%, a few not covered due to sequence complexity) for a total of 0.067 Mb in 1236 amplicons. The OvCa panel covers 25 CDS and an additional 130 regions of interest, with an average target coverage for the 0.114 Mb of 99.25% in 1672 amplicons.

### Supplementary Note 2

To decipher tumor composition, markers and expression signatures for tumor cells themselves (e.g., MelA, MITF, SOX9 for melanoma; CD44, CD133, CD24 for ovarian cancer; and cyMPO, CD117, CD14 in AML), immune cell subtypes (such as CD8, CD68, and CD20), endothelial cells (CD31) and fibroblasts (SMA, FAP) are used in the CyTOF, scRNA, digital pathology and proteomics technologies. The scDNA technology bases tumor cell identification on overall copy number variation profiles in tumor cells versus tumor-associated cells, whereas the drug profiling technologies use a reduced set of markers to differentiate cancer from immune cells.

To provide clinical decision support, a panel of small molecule and chemotherapy drugs including their prioritisation for the drug profiling technologies, for each one of the TuPro indications was created. These panels include approved drugs for the respective indication (such as BRAF inhibitors for melanoma and the protein kinase inhibitor midostaurin in AML) as well as off-label drugs that target the most common genetic aberrations in the respective tumor types (such as CDK4/6 inhibitors in melanoma targeting *CDK4* amplification and IDH1 inhibitors in AML targeting activating *IDH1* mutations). Off-label therapeutics that have been shown to induce responses in the respective preclinical models and/or are in clinical trials in Switzerland for the indication (e.g., the aromatase inhibitor letrozole for ovarian cancer, and ERK inhibitors for melanoma) are also included. Based on this small molecule and chemotherapy drug list as well as currently available (approved or in trials) immunotherapies, the corresponding panels of treatment-relevant markers or genes of interest for the single technologies were developed. The panels include markers relevant for immunotherapy, such as PD-L1, TIM-3, MHC class I, and markers relevant for therapies targeted towards oncogenic pathways, such as pEGFR, pMET, pERK. During the course of TuPro, with new predictive biomarkers being discovered frequently, the marker panels may evolve and get updated. Importantly, although these lists are designed for clinical decision support, the non-targeted technologies such as proteomics, scRNA, and bulk RNA carry out non-targeted biomarker discovery.

## TUPRO Consortium

Rudolf Aebersold (2), Faisal S Al-Quaddoomi (8,15), Jonas Albinus (7), Ilaria Alborelli (23), Per-Olof Attinger (10), Marina Bacac (14), Daniel Baumhoer (23), Beatrice Beck-Schimmer (36), Niko Beerenwinkel (4), Christian Beisel (4), Lara Bernasconi (26), Anne Bertolini (8,15), Bernd Bodenmiller (32), Ximena Bonilla (3,6,15,25), Ruben Casanova (32), Stéphane Chevrier (32), Natalia Chicherova (8,15), Maya D’Costa (9), Esther Danenberg (34), Natalie Davidson (3,6,15,25), Reinhard Dummer (27), Stefanie Engler (32), Martin Erkens (12), Katja Eschbach (4), Cinzia Esposito (34), André Fedier (16), Pedro Ferreira (4), Joanna Ficek (3,6,15,25), Anja L Frei (28), Bruno Frey (11), Sandra Goetze (7), Linda Grob (8,15), Detlef Günther (5), Gabriele Gut (34), Pirmin Haeuptle (1), Viola Heinzelmann-Schwarz (16,22), Sylvia Herter (14), Rene Holtackers (34), Tamara Huesser (14), Anja Irmisch (27), Francis Jacob (16), Andrea Jacobs (32), Tim M Jaeger (10), Katharina Jahn (4), Alva R James (3,6,15,25), Philip M Jermann (23), André Kahles (3,6,15,25), Abdullah Kahraman (15,28), Viktor H Koelzer (28), Werner Kuebler (24), Jack Kuipers (4), Christian P Kunze (21), Christian Kurzeder (19), Jelena Kühn-Georgijevic (12), Kjong-Van Lehmann (3,6,15,25), Mitchell Levesque (27), Sebastian Lugert (9), Gerd Maass (11), Sergio Maffioletti (33), Julien Mena (2), Ulrike Menzel (4), Nicola Miglino (1), Emanuela S Milani (7), Holger Moch (28), Simone Muenst (23), Riccardo Murri (35), Charlotte KY Ng (23,31), Stefan Nicolet (23), Patrick GA Pedrioli (2), Lucas Pelkmans (34), Salvatore Piscuoglio (16,23), Michael Prummer (8,15), Mathilde Ritter (16), Christian Rommel (12), María L Rosano-González (8,15), Gunnar Rätsch (3,6,15,25), Jacobo Sarabia del Castillo (34), Ramona Schlenker (13), Petra C Schwalie (12), Severin Schwan (10), Tobias Schär (4), Gabriela Senti (26), Franziska Singer (8,15), Berend Snijder (2), Bettina Sobottka (28), Vipin T Sreedharan (8,15), Stefan Stark (3,6,15,25), Daniel J Stekhoven (8,15), Tinu M Thomas (3,6,15,25), Markus Tolnay (23), Nora C Toussaint (8,15), Mustafa A Tuncel (4), Audrey Van Drogen (7), Marcus Vetter (18), Tatjana Vlajnic (23), Sandra Weber (26), Walter P Weber (17), Rebekka Wegmann (2), Michael Weller (30), Fabian Wendt (7), Norbert Wey (28), Andreas Wicki (1,16,20), Bernd Wollscheid (7), Shuqing Yu (8,15), Johanna Ziegler (27), Marc Zimmermann (3,6,15,25), Martin Zoche (28), Gregor Zuend (29)

(1) Cantonal Hospital Baselland, Medical University Clinic, Rheinstrasse 26, 4410 Liestal, Switzerland, (2) ETH Zurich, Department of Biology, Otto-Stern-Weg 3, 8093 Zurich, Switzerland, (3) ETH Zurich, Department of Biology, Wolfgang-Pauli-Strasse 27, 8093 Zurich, Switzerland, (4) ETH Zurich, Department of Biosystems Science and Engineering, Mattenstrasse 26, 4058 Basel, Switzerland, (5) ETH Zurich, Department of Chemistry and Applied Biosciences, Vladimir-Prelog-Weg 1-5/10, 8093 Zurich, Switzerland, (6) ETH Zurich, Department of Computer Science, Institute of Machine Learning, Universitätstrasse 6, 8092 Zurich, Switzerland, (7) ETH Zurich, Department of Health Sciences and Technology, Otto-Stern-Weg 3, 8093 Zurich, Switzerland, (8) ETH Zurich, NEXUS Personalized Health Technologies, John-von-Neumann-Weg 9, 8093 Zurich, Switzerland, (9) F. Hoffmann-La Roche Ltd, Grenzacherstrasse 124, 4070 Basel, Switzerland, (10) F. Hoffmann-La Roche Ltd, Grenzacherstrasse 124, 4070 Basel, Switzerland,, (11) Roche Diagnostics GmbH, Nonnenwald 2, 82377 Penzberg, Germany, (12) Roche Pharmaceutical Research and Early Development, Roche Innovation Center Basel, Grenzacherstrasse 124, 4070 Basel, Switzerland, (13) Roche Pharmaceutical Research and Early Development, Roche Innovation Center Munich, Roche Diagnostics GmbH, Nonnenwald 2, 82377 Penzberg, Germany, (14) Roche Pharmaceutical Research and Early Development, Roche Innovation Center Zurich, Wagistrasse 10, 8952 Schlieren, Switzerland, (15) Swiss Institute of Bioinformatics, Zurich, Switzerland, (16) University Hospital Basel and University of Basel, Department of Biomedicine, Hebelstrasse 20, 4031 Basel, Switzerland, (17) University Hospital Basel and University of Basel, Department of Surgery, Brustzentrum, Spitalstrasse 21, 4031 Basel, Switzerland, (18) University Hospital Basel, Brustzentrum & Tumorzentrum, Petersgraben 4, 4031 Basel, Switzerland, (19) University Hospital Basel, Brustzentrum, Spitalstrasse 21, 4031 Basel, Switzerland, (20) University Hospital Basel, Centre for Neuroendocrine & Endocrine Tumours, Spitalstrasse 21/Petersgraben 4, 4031 Basel, Switzerland, (21) University Hospital Basel, Department of Information- and Communication Technology, Spitalstrasse 26, 4031 Basel, Switzerland, (22) University Hospital Basel, Gynecological Cancer Center, Spitalstrasse 21, 4031 Basel, Switzerland, (23) University Hospital Basel, Institute of Medical Genetics and Pathology, Schönbeinstrasse 40, 4031 Basel, Switzerland, (24) University Hospital Basel, Spitalstrasse 21/Petersgraben 4, 4031 Basel, Switzerland, (25) University Hospital Zurich, Biomedical Informatics, Schmelzbergstrasse 26, 8006 Zurich, Switzerland, (26) University Hospital Zurich, Clinical Trials Center, Rämistrasse 100, 8091 Zurich, Switzerland, (27) University Hospital Zurich, Department of Dermatology, Gloriastrasse 31, 8091 Zurich, Switzerland, (28) University Hospital Zurich, Department of Pathology and Molecular Pathology, Schmelzbergstrasse 12, 8091 Zurich, Switzerland, (29) University Hospital Zurich, Rämistrasse 100, 8091 Zurich, Switzerland, (30) University Hospital and University of Zurich, Department of Neurology, Frauenklinikstrasse 26, 8091 Zurich, Switzerland, (31) University of Bern, Department of BioMedical Research, Murtenstrasse 35, 3008 Bern, Switzerland, (32) University of Zurich, Department of Quantitative Biomedicine, Winterthurerstrasse 190, 8057 Zurich, Switzerland, (33) University of Zurich, Grid Computing Competence Center, Rämistrasse 71, 8006 Zurich, Switzerland, (34) University of Zurich, Institute of Molecular Life Sciences, Winterthurerstrasse 190, 8057 Zurich, Switzerland, (35) University of Zurich, Services and Support for Science IT, Winterthurerstrasse 190, 8057 Zurich, Switzerland, (36) University of Zurich, VP Medicine, Künstlergasse 15, 8001 Zurich, Switzerland

## Notes

### Competing Interest Statement

The authors have declared no competing interest.

### Clinical Trial

BASEC-Nr.2018-02050

### Funding Statement

The study described in this paper is the result of a jointly-funded effort between several academic institutions (The University of Zurich, The University of Zurich Hospital, The Swiss Federal Institute of Technology in Zurich, The University of Basel Hospital, and The University of Basel), as well as F. Hoffmann-La Roche AG.

## References

Aebersold, R., and M. Mann. 2016. “Mass-Spectrometric Exploration of Proteome Structure and Function.” Nature. https://www.nature.com/articles/nature19949.

Bandura, Dmitry R., Vladimir I. Baranov, Olga I. Ornatsky, Alexei Antonov, Robert Kinach, Xudong Lou, Serguei Pavlov, Sergey Vorobiev, John E. Dick, and Scott D. Tanner. 2009. “Mass Cytometry: Technique for Real Time Single Cell Multitarget Immunoassay Based on Inductively Coupled Plasma Time-of-Flight Mass Spectrometry.” Analytical Chemistry 81 (16): 6813–22.

Battich, Nico, Thomas Stoeger, and Lucas Pelkmans. 2015. “Control of Transcript Variability in Single Mammalian Cells.” Cell 163 (7): 1596–1610.

Beishline, Kate, and Jane Azizkhan-Clifford. 2014. “Interplay between the Cell Cycle and Double-Strand Break Response in Mammalian Cells.” Methods in Molecular Biology 1170: 41–59.

Beltran, Himisha, Kenneth Eng, Juan Miguel Mosquera, Alexandros Sigaras, Alessandro Romanel, Hanna Rennert, Myriam Kossai, et al. 2015. “Whole-Exome Sequencing of Metastatic Cancer and Biomarkers of Treatment Response.” JAMA Oncology 1 (4): 466–74.

Bettegowda, Chetan, Mark Sausen, Rebecca J. Leary, Isaac Kinde, Yuxuan Wang, Nishant Agrawal, Bjarne R. Bartlett, et al. 2014. “Detection of Circulating Tumor DNA in Early- and Late-Stage Human Malignancies.” Science Translational Medicine 6 (224): 224ra24.

Campbell, P. J., G. Getz, J. M. Stuart, J. O. Korbel, and L. D. Stein. 2017. “Pan-Cancer Analysis of Whole Genomes.” BioRxiv. https://www.biorxiv.org/content/10.1101/162784v1.abstract.

Cancer Genome Atlas Network. 2015. “Genomic Classification of Cutaneous Melanoma.” Cell 161 (7): 1681–96.

Chevrier, Stéphane, Jacob Harrison Levine, Vito Riccardo Tomaso Zanotelli, Karina Silina, Daniel Schulz, Marina Bacac, Carola Hermine Ries, et al. 2017. “An Immune Atlas of Clear Cell Renal Cell Carcinoma.” Cell 169 (4): 736–49.e18.

Cirenajwis, Helena, Martin Lauss, Henrik Ekedahl, Therese Törngren, Anders Kvist, Lao H. Saal, Håkan Olsson, et al. 2017. “NF1-Mutated Melanoma Tumors Harbor Distinct Clinical and Biological Characteristics.” Molecular Oncology 11 (4): 438–51.

Colombo, N., C. Sessa, A. du Bois, J. Ledermann, W. G. McCluggage, I. McNeish, P. Morice, et al. 2019. “ESMO–ESGO Consensus Conference Recommendations on Ovarian Cancer: Pathology and Molecular Biology, Early and Advanced Stages, Borderline Tumours and Recurrent Disease†.” Annals of Oncology. https://doi.org/10.1093/annonc/mdz062.

Damond, Nicolas, Stefanie Engler, Vito R. T. Zanotelli, Denis Schapiro, Clive H. Wasserfall, Irina Kusmartseva, Harry S. Nick, et al. 2019. “A Map of Human Type 1 Diabetes Progression by Imaging Mass Cytometry.” Cell Metabolism 29 (3): 755–68.e5.

Dummer, R., A. Hauschild, N. Lindenblatt, G. Pentheroudakis, and U. Keilholz. 2015. “Cutaneous Melanoma: ESMO Clinical Practice Guidelines for Diagnosis, Treatment and Follow-Up.” Annals of Oncology. https://doi.org/10.1093/annonc/mdv297.

Fey, M. F., C. Buske, and ESMO Guidelines Working Group. 2013. “Acute Myeloblastic Leukaemias in Adult Patients: ESMO Clinical Practice Guidelines for Diagnosis, Treatment and Follow-Up.” Annals of Oncology: Official Journal of the European Society for Medical Oncology / ESMO 24 Suppl 6 (October): vi138–43.

Frampton, Garrett M., Alex Fichtenholtz, Geoff A. Otto, Kai Wang, Sean R. Downing, Jie He, Michael Schnall-Levin, et al. 2013. “Development and Validation of a Clinical Cancer

Genomic Profiling Test Based on Massively Parallel DNA Sequencing.” Nature Biotechnology 31 (11): 1023–31.

Frei, Andreas P., Felice-Alessio Bava, Eli R. Zunder, Elena W. Y. Hsieh, Shih-Yu Chen, Garry P. Nolan, and Pier Federico Gherardini. 2016. “Highly Multiplexed Simultaneous Detection of RNAs and Proteins in Single Cells.” Nature Methods 13 (3): 269–75.

Friedman, Adam A., Anthony Letai, David E. Fisher, and Keith T. Flaherty. 2015. “Precision Medicine for Cancer with next-Generation Functional Diagnostics.” Nature Reviews. Cancer 15 (12): 747–56.

Giesen, Charlotte, Hao A. O. Wang, Denis Schapiro, Nevena Zivanovic, Andrea Jacobs, Bodo Hattendorf, Peter J. Schüffler, et al. 2014. “Highly Multiplexed Imaging of Tumor Tissues with Subcellular Resolution by Mass Cytometry.” Nature Methods 11 (4): 417–22.

Goel, Shom, Molly J. DeCristo, April C. Watt, Haley BrinJones, Jaclyn Sceneay, Ben B. Li, Naveed Khan, et al. 2017. “CDK4/6 Inhibition Triggers Anti-Tumour Immunity.” Nature 548 (7668): 471–75.

Guo, Tiannan, Petri Kouvonen, Ching Chiek Koh, Ludovic C. Gillet, Witold E. Wolski, Hannes L. Röst, George Rosenberger, et al. 2015. “Rapid Mass Spectrometric Conversion of Tissue Biopsy Samples into Permanent Quantitative Digital Proteome Maps.” Nature Medicine 21 (4): 407–13.

Gut, Gabriele, Markus D. Herrmann, and Lucas Pelkmans. 2018. “Multiplexed Protein Maps Link Subcellular Organization to Cellular States.” Science 361 (6401). https://doi.org/10.1126/science.aar7042.

Hedlund, Eva, and Qiaolin Deng. 2018. “Single-Cell RNA Sequencing: Technical Advancements and Biological Applications.” Molecular Aspects of Medicine 59 (February): 36–46.

Ho, Yu-Jui, Naishitha Anaparthy, David Molik, Grinu Mathew, Toby Aicher, Ami Patel, James Hicks, and Molly Gale Hammell. 2018. “Single-Cell RNA-Seq Analysis Identifies Markers of Resistance to Targeted BRAF Inhibitors in Melanoma Cell Populations.” Genome Research 28 (9): 1353–63.

Iontorret, Application Note. n.d. “Next-Generation Sequencing Analysis of Low-Frequency Mutations from Cell-Free DNA.” https://tools.thermofisher.com/content/sfs/brochures/ngs-analysis-mutations-cfdna-app-note.pdf.

Jahn, Katharina, Jack Kuipers, and Niko Beerenwinkel. 2016. “Tree Inference for Single-Cell Data.” Genome Biology 17 (May): 86.

Jayawardana, Kaushala, Sarah-Jane Schramm, Lauren Haydu, John F. Thompson, Richard A. Scolyer, Graham J. Mann, Samuel Müller, and Jean Yee Hwa Yang. 2015. “Determination of Prognosis in Metastatic Melanoma through Integration of Clinico-Pathologic, Mutation, mRNA, microRNA, and Protein Information.” International Journal of Cancer. Journal International Du Cancer 136 (4): 863–74.

Kahles, André, Kjong-Van Lehmann, Nora C. Toussaint, Matthias Hüser, Stefan G. Stark, Timo Sachsenberg, Oliver Stegle, et al. 2018. “Comprehensive Analysis of Alternative Splicing Across Tumors from 8,705 Patients.” Cancer Cell 34 (2): 211–24.e6.

Kuipers, Jack, Katharina Jahn, Benjamin J. Raphael, and Niko Beerenwinkel. 2017. “Single-Cell Sequencing Data Reveal Widespread Recurrence and Loss of Mutational Hits in the Life Histories of Tumors.” Genome Research 27 (11): 1885–94.

Lavin, Yonit, Soma Kobayashi, Andrew Leader, El-Ad David Amir, Naama Elefant, Camille Bigenwald, Romain Remark, et al. 2017. “Innate Immune Landscape in Early Lung Adenocarcinoma by Paired Single-Cell Analyses.” Cell 169 (4): 750–65.e17.

Lehmann, Kjong-Van, André Kahles, Cyriac Kandoth, William Lee, Nikolaus Schultz, Oliver Stegle, and Gunnar Rätsch. 2015. “Integrative Genome-Wide Analysis of the Determinants of RNA Splicing in Kidney Renal Clear Cell Carcinoma.” Pacific Symposium on Biocomputing. Pacific Symposium on Biocomputing, 44–55.

Le Tourneau, Christophe, Jean-Pierre Delord, Anthony Gonçalves, Céline Gavoille, Coraline Dubot, Nicolas Isambert, Mario Campone, et al. 2015. “Molecularly Targeted Therapy Based on Tumour Molecular Profiling versus Conventional Therapy for Advanced Cancer (SHIVA): A Multicentre, Open-Label, Proof-of-Concept, Randomised, Controlled Phase 2 Trial.” The Lancet Oncology 16 (13): 1324–34.

Massard, Christophe, Stefan Michiels, Charles Ferté, Marie-Cécile Le Deley, Ludovic Lacroix, Antoine Hollebecque, Loic Verlingue, et al. 2017. “High-Throughput Genomics and Clinical Outcome in Hard-to-Treat Advanced Cancers: Results of the MOSCATO 01 Trial.” Cancer Discovery 7 (6): 586–95.

Mateo, J., D. Chakravarty, R. Dienstmann, S. Jezdic, A. Gonzalez-Perez, N. Lopez-Bigas, C. K. Y. Ng, et al. 2018. “A Framework to Rank Genomic Alterations as Targets for Cancer Precision Medicine: The ESMO Scale for Clinical Actionability of Molecular Targets (ESCAT).” Annals of Oncology: Official Journal of the European Society for Medical Oncology / ESMO 29 (9): 1895–1902.

Meric-Bernstam, Funda, Lauren Brusco, Kenna Shaw, Chacha Horombe, Scott Kopetz, Michael A. Davies, Mark Routbort, et al. 2015. “Feasibility of Large-Scale Genomic Testing to Facilitate Enrollment Onto Genomically Matched Clinical Trials.” Journal of Clinical Oncology: Official Journal of the American Society of Clinical Oncology 33 (25): 2753–62.

Michels, J., I. Vitale, M. Saparbaev, M. Castedo, and G. Kroemer. 2014. “Predictive Biomarkers for Cancer Therapy with PARP Inhibitors.” Oncogene 33 (30): 3894–3907.

Navin, Nicholas, Jude Kendall, Jennifer Troge, Peter Andrews, Linda Rodgers, Jeanne McIndoo, Kerry Cook, et al. 2011. “Tumour Evolution Inferred by Single-Cell Sequencing.” Nature 472 (7341): 90–94.

Papalexi, Efthymia, and Rahul Satija. 2018. “Single-Cell RNA Sequencing to Explore Immune Cell Heterogeneity.” Nature Reviews. Immunology 18 (1): 35–45.

Paulson, K. G., V. Voillet, M. S. McAfee, D. S. Hunter, F. D. Wagener, M. Perdicchio, W. J. Valente, et al. 2018. “Acquired Cancer Resistance to Combination Immunotherapy from Transcriptional Loss of Class I HLA.” Nature Communications 9 (1): 3868.

PCAWG Transcriptome Core Group, Claudia Calabrese, Natalie R. Davidson, Nuno A. Fonseca, Yao He, André Kahles, Kjong-Van Lehmann, et al. 2018. “Genomic Basis for RNA Alterations Revealed by Whole-Genome Analyses of 27 Cancer Types.” bioRxiv. https://doi.org/10.1101/183889.

Peterson, Amelia C., Jason D. Russell, Derek J. Bailey, Michael S. Westphall, and Joshua J. Coon. 2012. “Parallel Reaction Monitoring for High Resolution and High Mass Accuracy Quantitative, Targeted Proteomics.” Molecular & Cellular Proteomics: MCP 11 (11): 1475–88.

Robinson, Dan R., Yi-Mi Wu, Robert J. Lonigro, Pankaj Vats, Erin Cobain, Jessica Everett, Xuhong Cao, et al. 2017. “Integrative Clinical Genomics of Metastatic Cancer.” Nature 548 (7667): 297–303.

Rodon, Jordi, Jean-Charles Soria, Raanan Berger, Wilson H. Miller, Eitan Rubin, Aleksandra Kugel, Apostolia Tsimberidou, et al. 2019. “Genomic and Transcriptomic Profiling Expands Precision Cancer Medicine: The WINTHER Trial.” Nature Medicine 25 (5): 751–58.

Röst, Hannes L., Lars Malmström, and Ruedi Aebersold. 2015. “Reproducible Quantitative Proteotype Data Matrices for Systems Biology.” Molecular Biology of the Cell 26 (22): 3926–31.

Schapiro, Denis, Hartland W. Jackson, Swetha Raghuraman, Jana R. Fischer, Vito R. T. Zanotelli, Daniel Schulz, Charlotte Giesen, Raúl Catena, Zsuzsanna Varga, and Bernd Bodenmiller. 2017. “histoCAT: Analysis of Cell Phenotypes and Interactions in Multiplex Image Cytometry Data.” Nature Methods 14 (9): 873–76.

Schiess, Ralph, Bernd Wollscheid, and Ruedi Aebersold. 2009. “Targeted Proteomic Strategy for Clinical Biomarker Discovery.” Molecular Oncology 3 (1): 33–44.

Schulz, Daniel, Vito Riccardo Tomaso Zanotelli, Jana Raja Fischer, Denis Schapiro, Stefanie Engler, Xiao-Kang Lun, Hartland Warren Jackson, and Bernd Bodenmiller. 2018. “Simultaneous Multiplexed Imaging of mRNA and Proteins with Subcellular Resolution in Breast Cancer Tissue Samples by Mass Cytometry.” Cell Systems 6 (1): 25–36.e5.

Shih, Andrew J., Andrew Menzin, Jill Whyte, John Lovecchio, Anthony Liew, Houman Khalili, Tawfiqul Bhuiya, Peter K. Gregersen, and Annette T. Lee. 2018. “Identification of Grade and Origin Specific Cell Populations in Serous Epithelial Ovarian Cancer by Single Cell RNA-Seq.” PloS One 13 (11): e0206785.

Sicklick, Jason K., Shumei Kato, Ryosuke Okamura, Maria Schwaederle, Michael E. Hahn, Casey B. Williams, Pradip De, et al. 2019. “Molecular Profiling of Cancer Patients Enables Personalized Combination Therapy: The I-PREDICT Study.” Nature Medicine 25 (5): 744–50.

Singer, Franziska, Anja Irmisch, Nora C. Toussaint, Linda Grob, Jochen Singer, Thomas Thurnherr, Niko Beerenwinkel, et al. 2018. “SwissMTB: Establishing Comprehensive Molecular Cancer Diagnostics in Swiss Clinics.” BMC Medical Informatics and Decision Making 18 (1): 89.

Singer, Jochen, Jack Kuipers, Katharina Jahn, and Niko Beerenwinkel. 2018. “Single-Cell Mutation Identification via Phylogenetic Inference.” Nature Communications 9 (1): 5144.

Snijder, Berend, Raphael Sacher, Pauli Rämö, Eva-Maria Damm, Prisca Liberali, and Lucas Pelkmans. 2009. “Population Context Determines Cell-to-Cell Variability in Endocytosis and Virus Infection.” Nature 461 (7263): 520–23.

Snijder, Berend, Gregory I. Vladimer, Nikolaus Krall, Katsuhiro Miura, Ann-Sofie Schmolke, Christoph Kornauth, Oscar Lopez de la Fuente, et al. 2017. “Image-Based Ex-Vivo Drug Screening for Patients with Aggressive Haematological Malignancies: Interim Results from a Single-Arm, Open-Label, Pilot Study.” The Lancet. Haematology 4 (12): e595–606.

Stewart, Caitlin M., Prachi D. Kothari, Florent Mouliere, Richard Mair, Saira Somnay, Ryma Benayed, Ahmet Zehir, et al. 2018. “The Value of Cell-Free DNA for Molecular Pathology.” The Journal of Pathology 244 (5): 616–27.

Svensson, Valentine, Roser Vento-Tormo, and Sarah A. Teichmann. 2018. “Exponential Scaling of Single-Cell RNA-Seq in the Past Decade.” Nature Protocols 13 (4): 599–604.

The GTEx Consortium. 2015. “The Genotype-Tissue Expression (GTEx) Pilot Analysis: Multitissue Gene Regulation in Humans.” Science 348 (6235): 648–60.

Tirosh, Itay, Benjamin Izar, Sanjay M. Prakadan, Marc H. Wadsworth 2nd, Daniel Treacy, John J. Trombetta, Asaf Rotem, et al. 2016. “Dissecting the Multicellular Ecosystem of Metastatic Melanoma by Single-Cell RNA-Seq.” Science 352 (6282): 189–96.

Wagner, Johanna, Maria Anna Rapsomaniki, Stéphane Chevrier, Tobias Anzeneder, Claus Langwieder, August Dykgers, Martin Rees, et al. 2019. “A Single-Cell Atlas of the Tumor and Immune Ecosystem of Human Breast Cancer.” Cell 177 (5): 1330–45.e18.

Yanez, B., T. Pearman, C. G. Lis, J. L. Beaumont, and D. Cella. 2013. “The FACT-G7: A Rapid Version of the Functional Assessment of Cancer Therapy-General (FACT-G) for Monitoring Symptoms and Concerns in Oncology Practice and Research.” Annals of Oncology: Official Journal of the European Society for Medical Oncology / ESMO 24 (4): 1073–78.

Zahn, Hans, Adi Steif, Emma Laks, Peter Eirew, Michael VanInsberghe, Sohrab P. Shah, Samuel Aparicio, and Carl L. Hansen. 2017. “Scalable Whole-Genome Single-Cell Library Preparation without Preamplification.” Nature Methods 14 (2): 167–73.

